# What is the extent of research assessing patients’ and clinicians’ perspectives on clozapine treatment? – a comprehensive scoping review

**DOI:** 10.1101/2024.02.29.24303563

**Authors:** Michelle Iris Jakobsen, Julie Perrine Schaug, Ole Jakob Storebø, Stephen Fitzgerald Austin, Jimmi Nielsen, Erik Simonsen

**Author notes:** **Correspondence to:** Michelle Iris Jakobsen, Psychiatric Research Unit East, The Mental Health Services of Region Zealand Psychiatry East, Smedegade 16, 4000 Roskilde, Denmark.

## Abstract

**Background:** The atypical antipsychotic clozapine is the gold standard for treating treatment-resistant schizophrenia; however, it is continuously underutilized in most parts of the world.

A few systematic reviews addressing barriers to clozapine prescribing have previously been conducted, primarily focusing on clinical staff’s attitudes and perceived barriers to prescribing. However, a preliminary literature search revealed that additional literature on the subject does exist, including literature on patient perspectives, without having been included in any of the former reviews.

It is therefore difficult to conclude if the former synthesizes of the literature are representative of current evidence, and if the topic has been adequately investigated to inform clinical practice. A scoping review is warranted to map and synthesize a broader scope of primary studies on patients’ and/or clinicians’ perspectives on clozapine treatment to identify any gaps for future research.

**Methods:** The electronic databases Cochrane Library, CINAHL, Web of Science, Psychinfo, MEDLINE, and EMBASE were searched, supplied with searches of Google Scholar, The Networked Digital Library of Theses and Dissertations (NDLTD), and OpenGrey. Citation tracking of selected studies was furthermore undertaken. Two researchers independently screened and extracted the data.

**Registration:** PROSPERO does not offer registration of scoping reviews; however, the protocol was prospectively registered with the Open Science Framework and subsequently published as an article.

**Results:** One hundred and forty-six studies were included. Most studies reported upon patients’ or clinicians’ perspectives on active clozapine treatment or on clinicians’ general perspectives on barriers to clozapine initiation. Three apparent gaps in research were identified: 1) clozapine eligible, yet clozapine-naïve, outpatients’ attitudes towards clozapine commencement, 2) assessments of clinicians’ reasons for clozapine withholding and perceived facilitators of clozapine treatment in specific patient-cases, and 3) direct assessments of both patient and clinician perspectives on clozapine discontinuation, continuation and re-challenge in specific patient-cases.

**Conclusions:** Research regarding perspectives on clozapine treatment tends to repeat itself. Future studies addressing the identified gaps in evidence are warranted and could provide the insights needed to optimize clozapine utilization.

**Strengths and limitations of this study:**

- The prospective registration and publication of the review protocol has ensured transparency of the review process.
- The search strategy has ensured a comprehensive search of the literature and multiple booster searches on Google Scholar have ensured a continued update on the scope of literature, the most recent one in January 2024.
- The original literature search was conducted in June 2021.
- The search was restricted to publications in the English language, which may have precluded the identification of some relevant insights and studies.

## 1. Introduction

Schizophrenia is a serious mental illness with major societal, social, and personal costs ^1^. Early, adequate treatment is crucial in order to improve the long-term outcome ^2 3^, however, approximately one-third of patients with schizophrenia fail to respond adequately to at least two conventional antipsychotics ^4 5^ and are considered treatment-resistant ^6^.

The antipsychotic (AP) clozapine has shown superior to other APs in terms of overall symptom reduction for schizophrenia and related disorders ^7^ and most guidelines recommend that clozapine should be offered first line to patients with treatment-resistant schizophrenia (TRS) ^6 8 9^.

It is furthermore indicated for a range of other disorders dependent on national guidelines^9^, e.g. schizophrenia or schizoaffective disorder with recurrent suicidality, treatment-resistant schizoaffective or bipolar disorder, psychotic disorders with poor tolerance to conventional neuroleptics, treatment-resistant organic psychosis, etc. Despite the recommendations and established advantages of clozapine treatment, clozapine is underutilized in most parts of the world ^10–15^, as it has been for decades, despite a steady flow of studies on the subject^16^.

The clozapine underutilization represents a major mental health concern and a few systematic reviews ^17–21^ have aimed to summarize the identified barriers and facilitators of clozapine prescribing. These previous reviews tend to include the same studies, primarily surveys of clinical staff’s attitudes and perceived barriers to prescribing and until recently, only a few studies examining patient perspectives on clozapine treatment^22–25^, have been included^17 18 20^. However, since the publication of the protocol article for this study^26^, two systematic reviews on patient perspectives have been published ^27 28^, reporting on a further sixteen studies^29–44^.

Nevertheless, our preliminary literature search revealed that additional studies on both patients’ and clinicians’ perspectives on clozapine treatment do exist, without having been included in any of the existing systematic reviews.

Consequently, it is difficult to conclude if the topic has been adequately elucidated to inform clinical practice, or if the lack of change in clozapine prescribing might be due to an inadequate dissemination and/or implementation of current evidence. A broader and more comprehensive overview of the literature addressing patients’ and/or clinicians’ perspectives on clozapine treatment is therefore warranted.

### 1.1. Key definitions

#### Patients

The term “patients” refers to adult (age ≥18 years) patients affiliated with somatic or mental health services.

#### Clinicians

The term “clinicians” will be used for all clinical staff affiliated with somatic or mental health services treating adult patients.

### 1.2. Objectives

In line with the original framework by Arksey and O’Malley ^45^ we aimed to conduct a scoping review in order to a) investigate the extent and variety of primary studies covering patients’ and/or clinicians’ perspectives of clozapine treatment, and b) to identify gaps in the current research. A secondary aim was to c) summarize the key findings related to patients’ and clinicians’ perspectives on clozapine treatment.

## 2. Methods and analysis

The protocol for this scoping review^26^ was designed in concordance with the Preferred Reporting Items for Systematic reviews and Meta-Analyses extension for Scoping Reviews (PRISMA-ScR) ^46^ to ensure that all relevant information for the future scoping review could be included. It was guided by the corresponding Joanna Briggs Institute (JBI) guidelines “Guidance for conducting systematic scoping reviews” ^47^ and the “Updated methodological guidance for the conduct of scoping reviews” ^48^. We have furthermore sought complemental guidance in the advanced framework recommendations by Levac et al.^49^.

A completed PRISMA checklist for the reporting of scoping reviews (Supplementary file 1 (S1)) has been submitted with the manuscript.

### 2.1. Registration

PROSPERO does not offer registration of scoping reviews; however, the review protocol was prospectively registered with the Open Science Framework (OSF), registration DOI 10.17605/OSF.IO/5K4S3^50^, and subsequently published as an article^26^.

### 2.2. Eligibility criteria

Publications were considered eligible for inclusion if they met the following selection criteria:

#### Inclusion criteria

- Published in the English language
- Primary, empirical literature addressing patients’ or clinicians’ perspectives on clozapine treatment (including both peer-reviewed research papers and grey literature such as conference abstracts and dissertation papers)

#### Exclusion criteria

- Non-empirical literature (i.e. editorials, opinion- and discussion papers) and case reports
- Secondary studies (reviews/overviews); however, their reference lists were included for citation tracking.

#### Rationale

We chose to limit our search to empirical, primary studies. The rationale for this was our aim to map the scope of studies on perspectives/attitudes/perceptions, with the intent to identify gaps in the existing scientific evidence. However, in order to uphold a certain level of scientific evidence, we disregarded case reports as sources of evidence.

We considered non-empirical data and secondary studies irrelevant to the objective of this review.

Due to feasibility resources, the language is restricted to English. No limitation has been set for year of publication or type of study.

### 2.3. Information sources

The electronic databases Cochrane Library, CINAHL, Web of Science, Psychinfo, MEDLINE and EMBASE were searched for relevant publications, supplemented with a booster search on Google scholar. This combination of sources have previously been reported to guarantee adequate and efficient coverage ^51^.

Furthermore, The Networked Digital Library of Theses and Dissertations (NDLTD) and OpenGrey were searched for additional relevant literature, and citation tracking of selected studies was undertaken.

### 2.4. Search

The search strategy used for this scoping review was developed by the lead investigator (author MIJ) in collaboration with an experienced research librarian. In accordance with established scoping review methodology ^47 48^, it consisted of three steps: First, an initial search of selected databases, in this case MEDLINE and EMBASE, was performed. Search terms included, but were not restricted to, clinician, doctor, psychiatrist, patient, consumer, perspective, experience, attitude, perception, clozapine, (leponex) and (clozaril). The search was then followed by screening of the identified articles for relevant text words and index terms. Secondly, the search was refined, incorporating the identified keywords and index terms. Search terms were adapted to the requirements of each selected database. Table 1 shows the refined electronic search strategy for Embase (originally published with the scoping review protocol^26^).

**Table 1.** Electronic search strategy for Embase.

| Search # | Search details |
| --- | --- |
| 1 | clozapine/ |
| 2 | (clozapin* or denzapin* or zaponex* or clozaril* or clopin* or fazaclo* or versaclo* or leponex*).ab,ti,tn,tw. |
| 3 | 1 or 2 |
| 4 | attitude/ |
| 5 | attitude assessment/ |
| 6 | exp satisfaction/ |
| 7 | (attitude* or belief* or perception* or view* or experience* or opinion* or perspective* or preference* or satisfaction or satisfied* or refus* or reason* or dislike* or content*).ab,ti,tw. |
| 8 | 4 or 5 or 6 or 7 |
| 9 | patient/ or mental patient/ or outpatient/ |
| 10 | consumer/ |
| 11 | (patient* or user* or consumer* or subject* or individual* or client*).ab,ti,tw. |
| 12 | physician/ |
| 13 | psychiatrist/ |
| 14 | clinician/ |
| 15 | (doctor* or physician* or psychiatrist* or clinician* or prescriber* or practitioner*).ab,ti,tw. |
| 16 | 9 or 10 or 11 or 12 or 13 or 14 or 15 |
| 17 | patient attitude/ or patient preference/ or patient satisfaction/ |
| 18 | physician attitude/ or health personnel attitude/ |
| 19 | consumer attitude/ |
| 20 | exp patient-reported outcome/ |
| 21 | exp treatment refusal/ |
| 22 | 17 or 18 or 19 or 20 or 21 |
| 23 | exp medication compliance/ or exp patient compliance/ |
| 24 | (barrier* or compliance*).ab,ti,tw. |
| 25 | 23 or 24 |
| 26 | 8 and 16 |
| 27 | 26 or 22 or 25 |
| 28 | 3 and 27 |
| 29 | limit 28 to english language |
| Notes<br>The electronic search strategy was adapted to the requirements of each selected database. The table shows the refined electronic search strategy for Embase and was first published with the protocol article<sup>26</sup>. |  |

After completing the refinement across all selected databases, a second search (the actual search), was undertaken by June 25, 2021. A Google Scholar booster search (screening only the first 200 hits and related titles) was undertaken by May 23, 2022. The third step of the search strategy consisted of searching the additional sources of literature (NDLTD and OpenGrey) on June 9, 2022, as well as hand-searches of the reference lists of all included studies and excluded review/overview studies, to ensure a comprehensive literature identification.

### 2.5. Selection of sources of evidence

Records from the database search and from additional sources were imported to the reference management software Endnote ^52^ and duplicates removed. The merged search results were then exported to Covidence ^53^. Two individual reviewers (authors MIJ and JPS) screened the titles and abstracts of all identified studies, followed by full-text screening according to the in- and exclusion criteria. Full texts were searched for, for all studies included for full-text screening. The search for full texts included assistance from a research librarian and requests for full texts on ResearchGate. If no full texts were available, studies were included for full-text screening in the form available. Any disagreement related to study eligibility at either stage was resolved through discussion between the two reviewers. Doubts regarding overall eligibility were solved by the involvement of a third party; e.g. we realized, during the screening phase, that some previous reviews/overviews had included studies reporting quantitative measures of well-being or quality of life. Through discussion with a third party (author OJS), we deemed such measures too influenced by other factors than the present use of antipsychotics and therefore not an assessment of perspectives on clozapine treatment. This decision led to the exclusion of studies reporting only these measures.

### 2.6. Data charting process

Microsoft® Excel ^54^, Covidence, and Endnote were used to manage the screening process and to organize the data.

A refined data extraction form, based on the preliminary form developed for the study protocol, was constructed in Covidence.

The preliminary data extraction form was refined to include pre-defined content categories (perspectives on active clozapine treatment/ discontinued clozapine treatment/ initiation of treatment) and sub-categories (e.g. satisfaction with / perceived burden of/ perceived efficacy of treatment, etc.) describing the perspectives being explored. The pre-defined categories and sub-categories were constructed based on the observations made during the screening process. The sub-category “Other” was added for additional contents not covered by the pre-defined sub-categories.

In accordance with the directed approach to content analysis^55^, the coding of study content was performed during the data extraction phase.

The content of “Other” was coded later on, during the analysis phase, to form additional sub-categories.

The extraction form was piloted twice, on ten studies, by both reviewers. After each pilot round, it was refined to record the information needed to answer the research questions. The data was extracted in doublet by the two reviewers and then combined into a consensus extraction. The lead investigator resolved any disagreements.

### 2.7. Data items

In the data charting phase of the review, the following information was collected:

- Author(s)
- Year of publication
- Title
- Country of origin
- Type of study
- Study population and study context/relevant characteristics (mixed staff/psychiatrists only, clozapine-/non-clozapine-/mixed patients, inpatients/outpatients etc.)
- Relevant outcome measures and method of assessment (any assessments of attitude or perception of clozapine treatment, e.g. measures of treatment satisfaction or efficacy, perception of barriers to its usage, reasons for treatment withholding or refusal, perceptions of initiatives for increased treatment utility, etc.)
- The content category(s) and sub-categories of explored perspectives (Perspectives related to active/discontinued/not yet commenced clozapine treatment? Perspectives related to specific patient cases or in general? Satisfaction with treatment?/ burden of treatment?/ efficacy of treatment?, etc.)
- Key findings related to patients’ or clinicians’ perspectives on clozapine treatment

### 2.8. Synthesis of results

A narrative summary of the results has been accompanied by visual aids. Studies were grouped and mapped according to the content categories of the explored perspectives (e.g. perspectives relating to active clozapine treatment/discontinued clozapine treatment/clozapine commencement etc.) and to the origin of perspectives (e.g. patients’/clinicians’ perspectives) to provide a graphic presentation of clusters and gaps in the existing evidence. A table presents an overview of basic study characteristics, while a supplementary spreadsheet presents an in-depth overview of individual study data (e.g. author, year, methods and key findings). Furthermore, simple column charts present distributions of explored sub-categories of perspectives. In line with the objectives of the review, recommendations for future investigations based on the identified pattern of previous research have been included in the conclusion.

### 2.9. Differences between the methods described in the published protocol and the methods used in the final review

In the protocol, we defined “patients” as adult patients affiliated with mental health services due to psychiatric disorders, while “clinicians” were defined as “psychiatrists” i.e. medical doctors affiliated with mental health services. However, during the screening process, we observed that several studies reported upon perspectives from mixed groups of patients, including patients with organic psychoses. These patients were sometimes, but not always, affiliated with mental health services which would entail an inconsistent exclusion of several sources of evidence. In the same way, we observed that several studies of clinician perspectives included mixed groups of clinicians, not all of them distinguishable in terms of psychiatrist vs non-psychiatrist perspectives, and not all of them including psychiatrists. Furthermore, psychiatrists are not the only prescribers of clozapine. Therefore, we changed the definitions to the current ones to ensure that all relevant studies were included for mapping.

## 3. Results

### 3.1. Selection of sources of evidence

On the basis of the initial search, 6531 studies were identified after duplicates were removed. At the title and abstract screening stage, 6160 studies were excluded. A total of 371 studies were screened as full texts (or at full-text level), and a further 225 studies were excluded for reasons outlined in the PRISMA flowchart, Figure 1.

**Figure 1.**
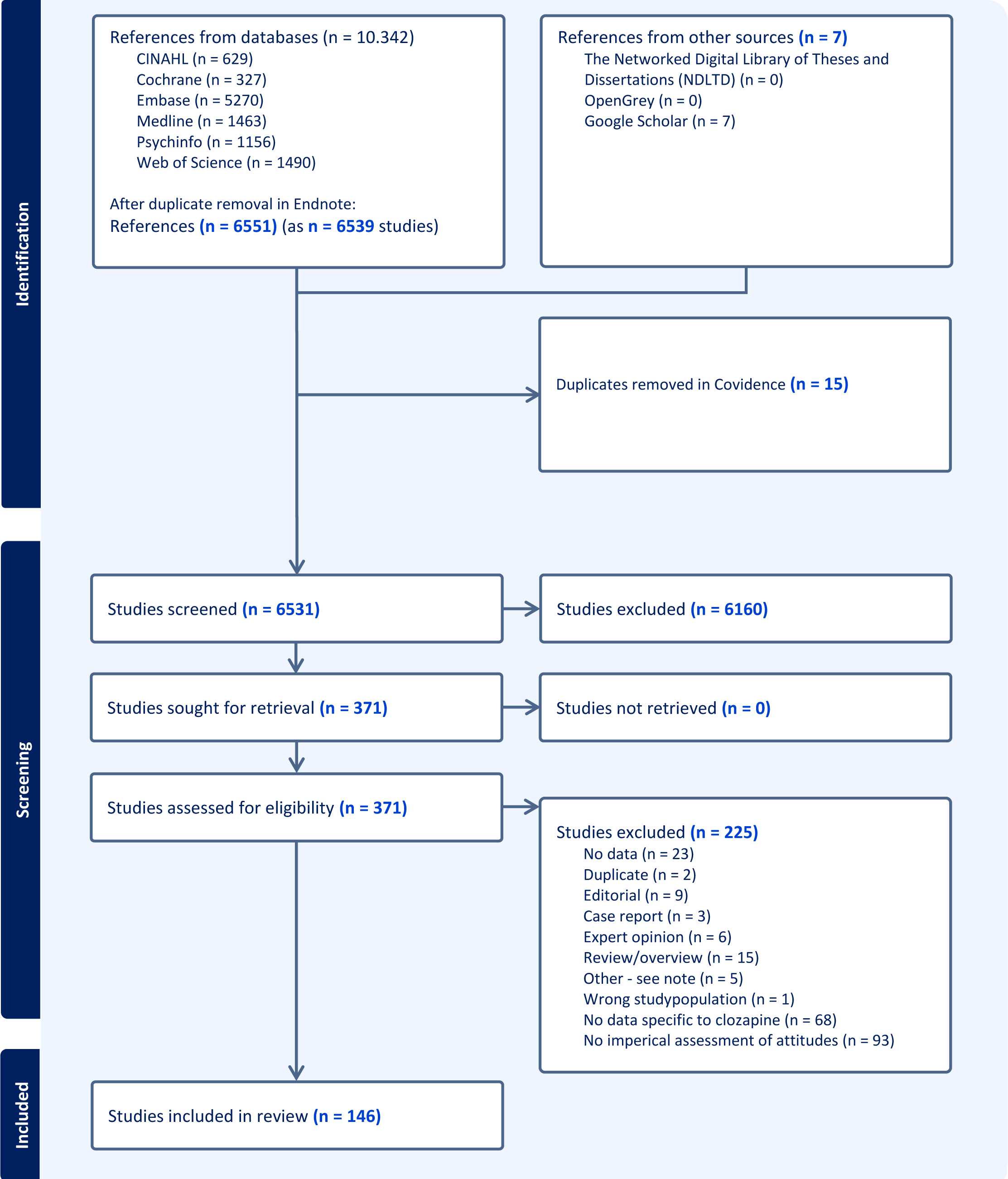
Screening. PRISMA flow-chart outlining the process of screening and inclusion of eligible sources of evidence.

### 3.2. General characteristics of included sources of evidence

Table 2 displays the individual study and participant characteristics.

**Table 2.**
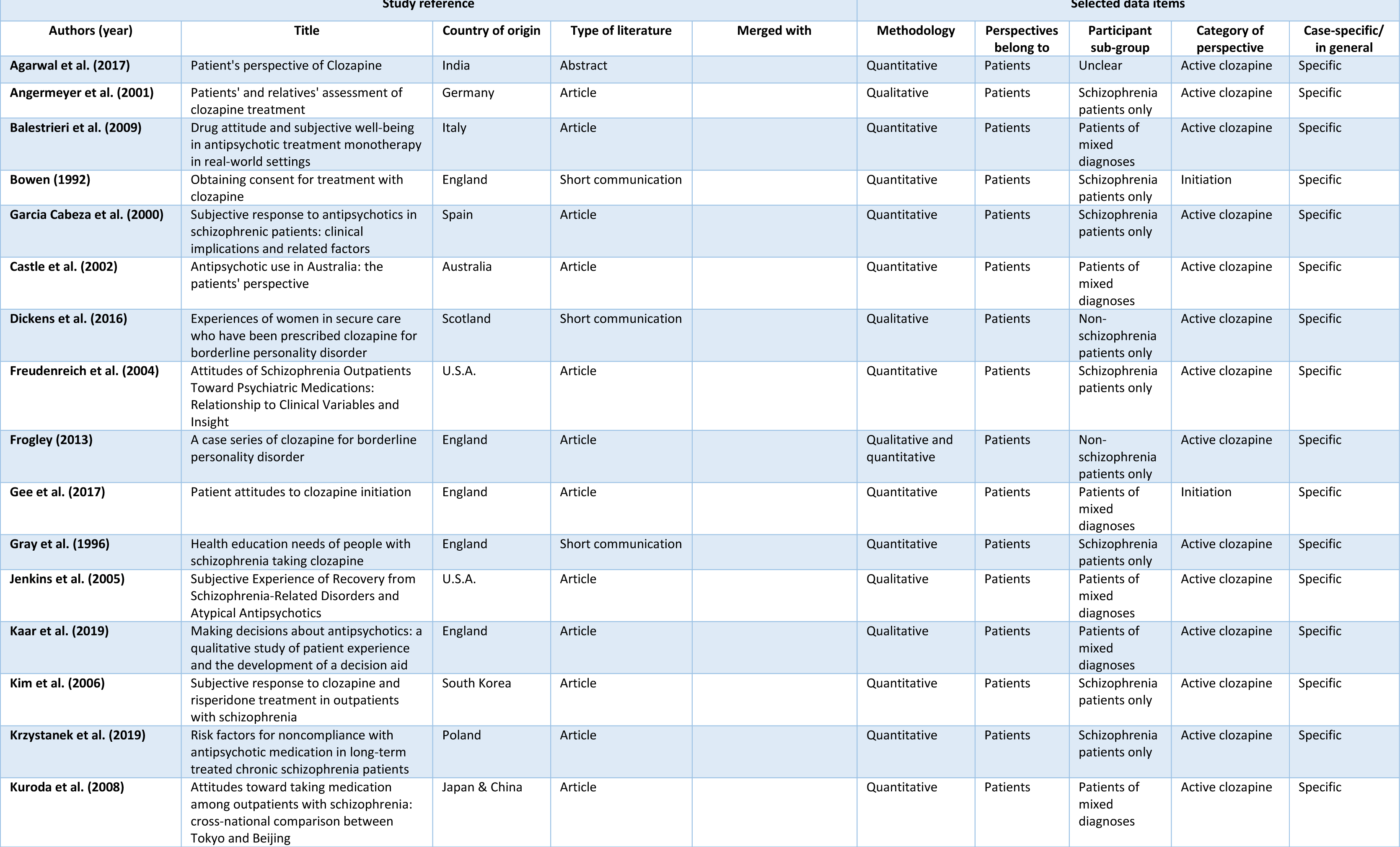
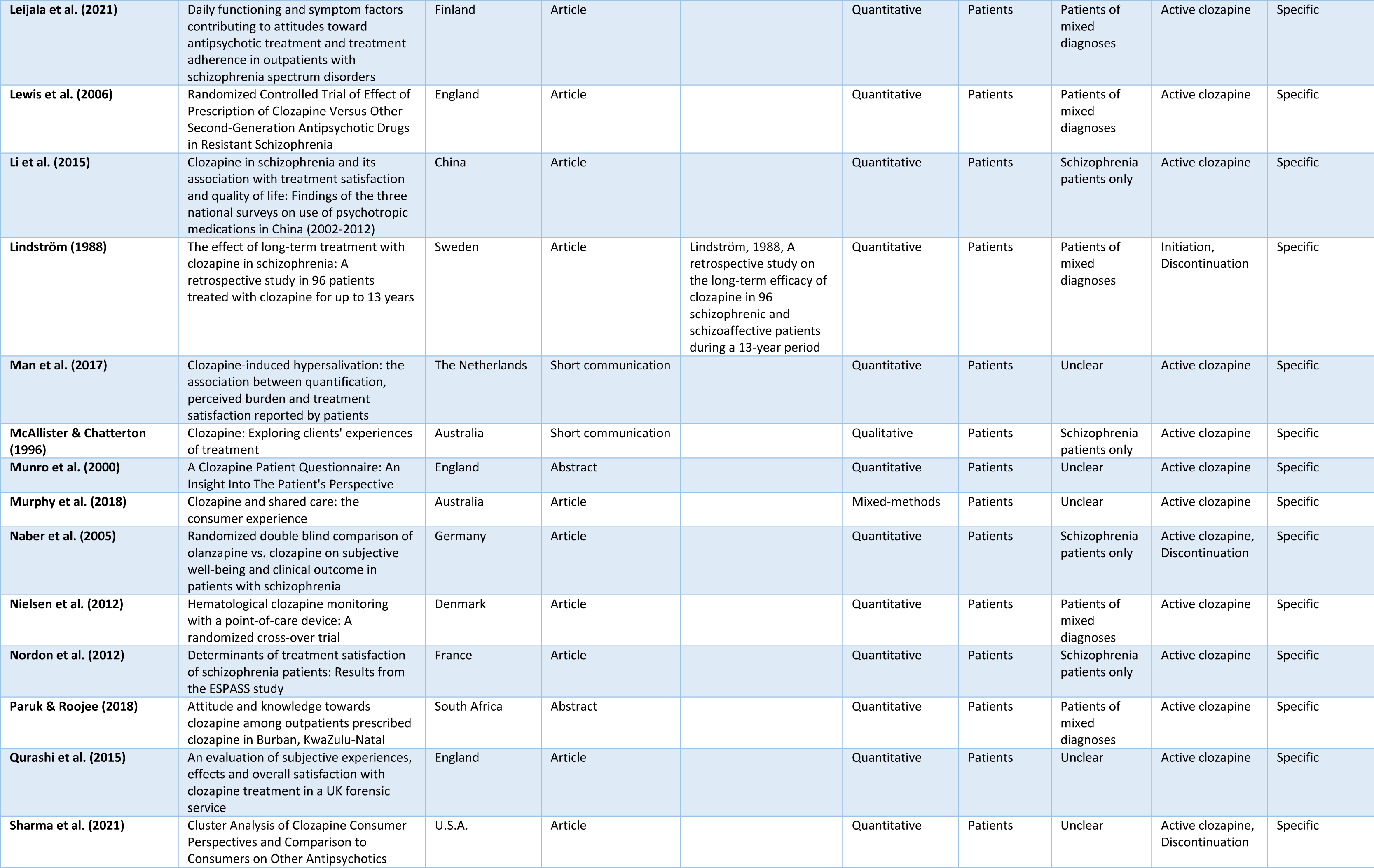
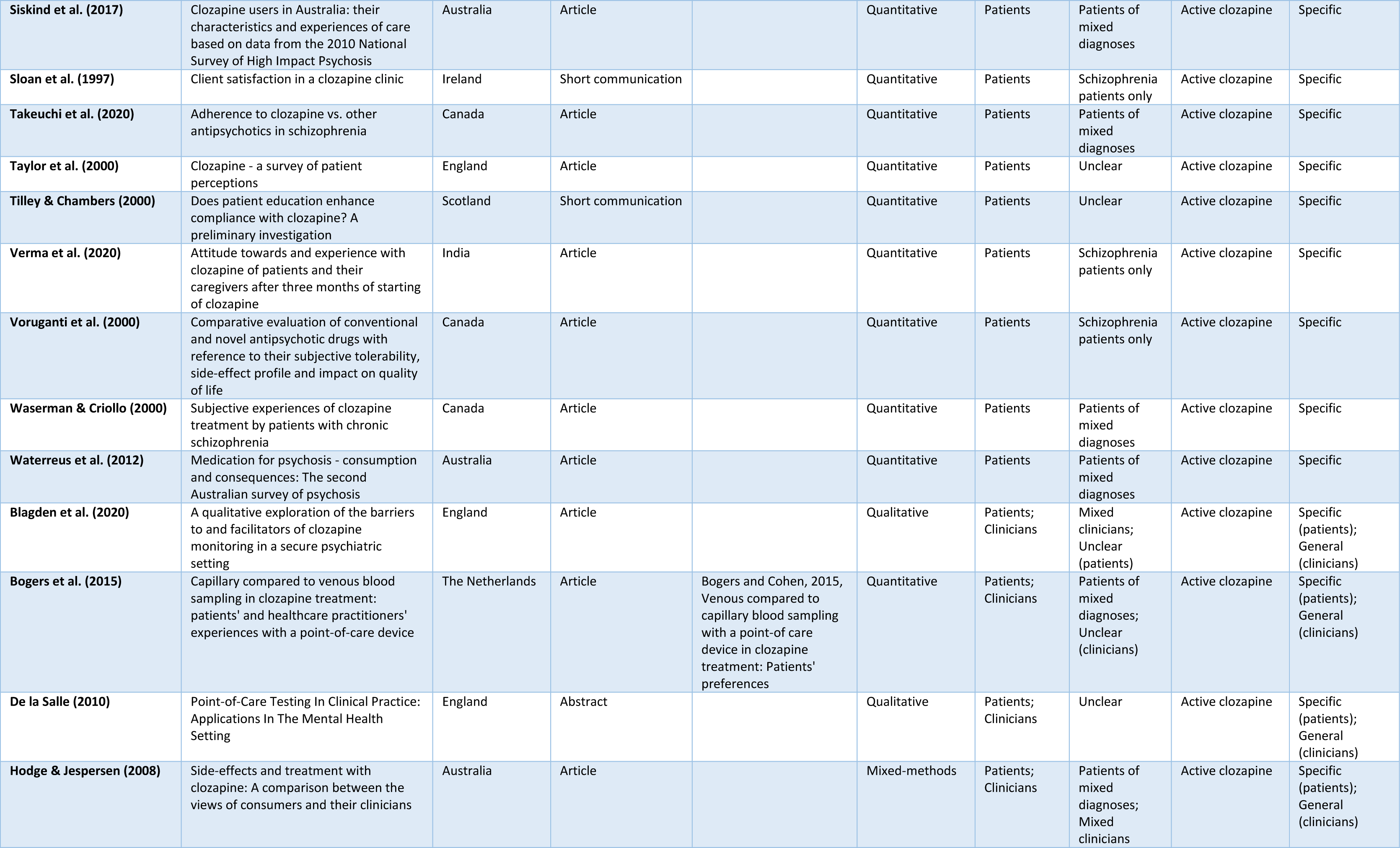
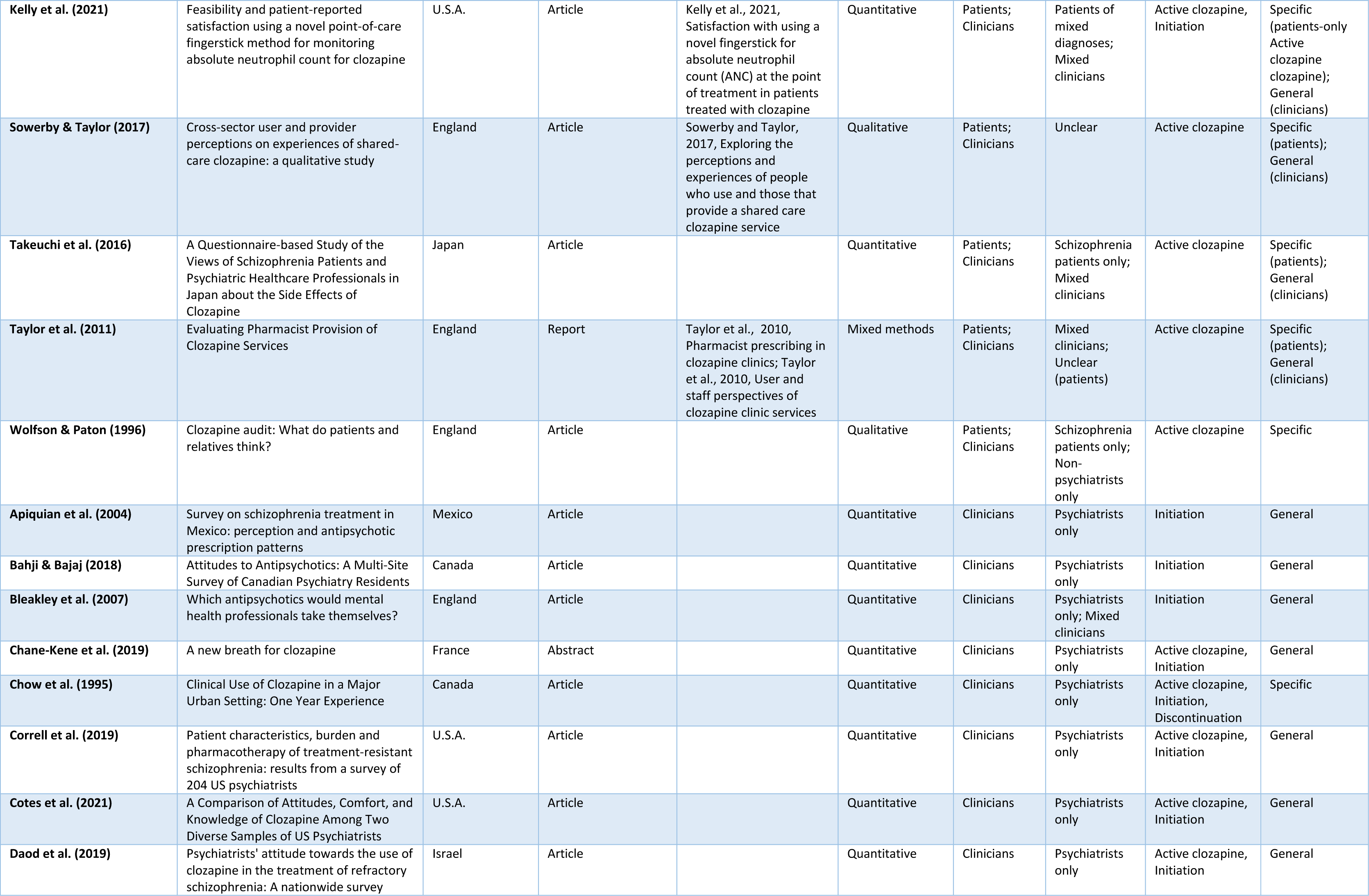
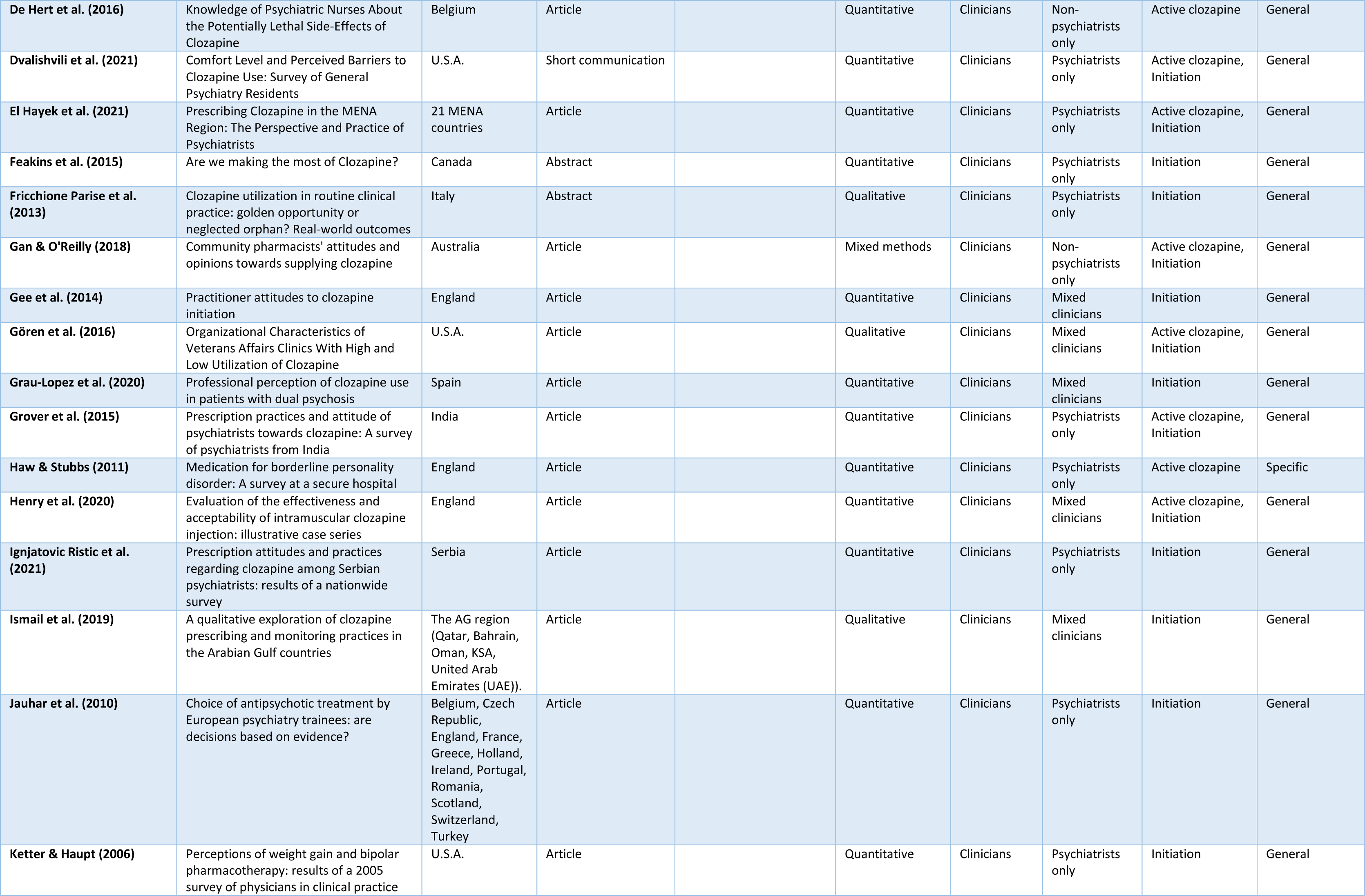
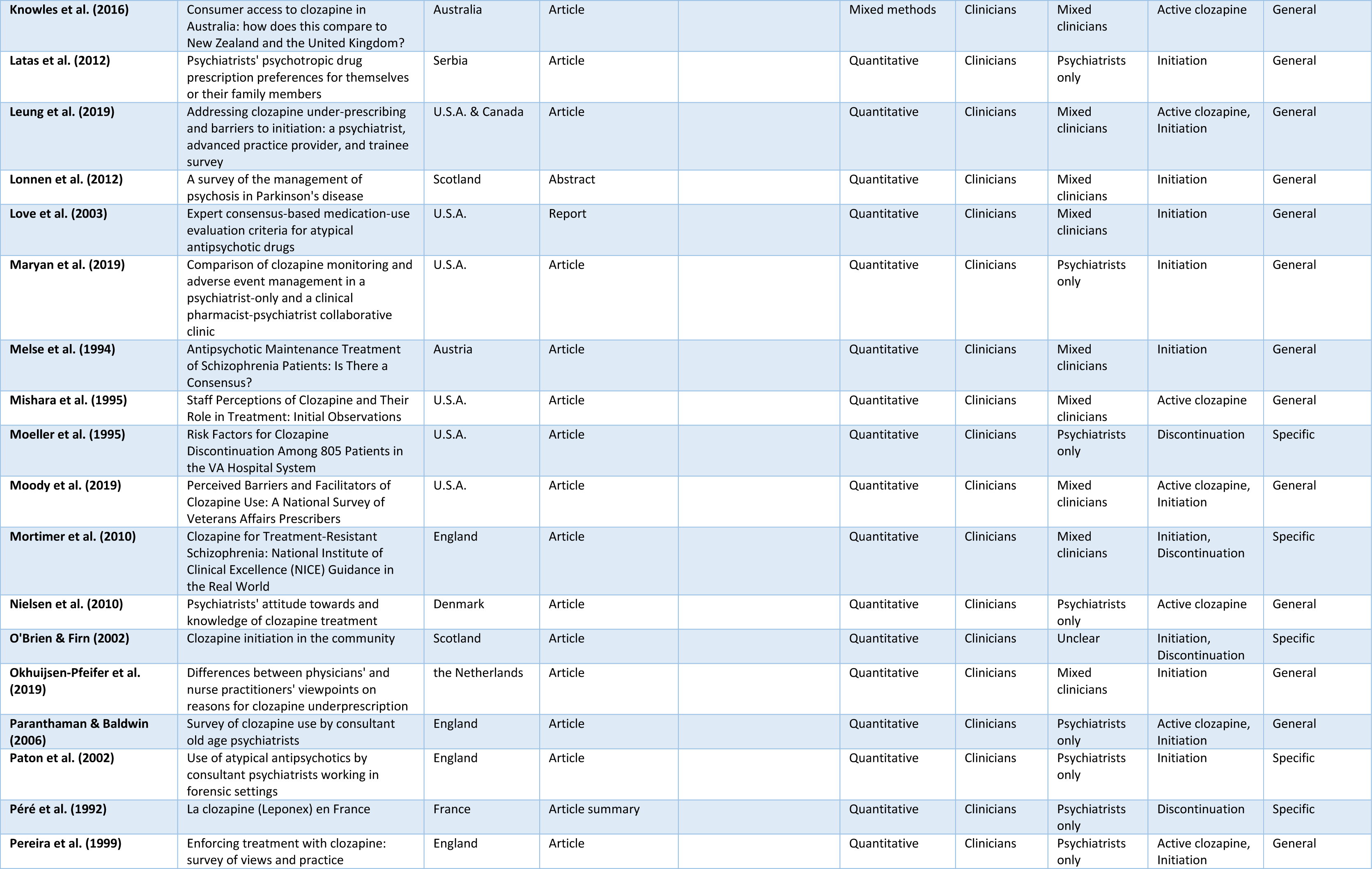
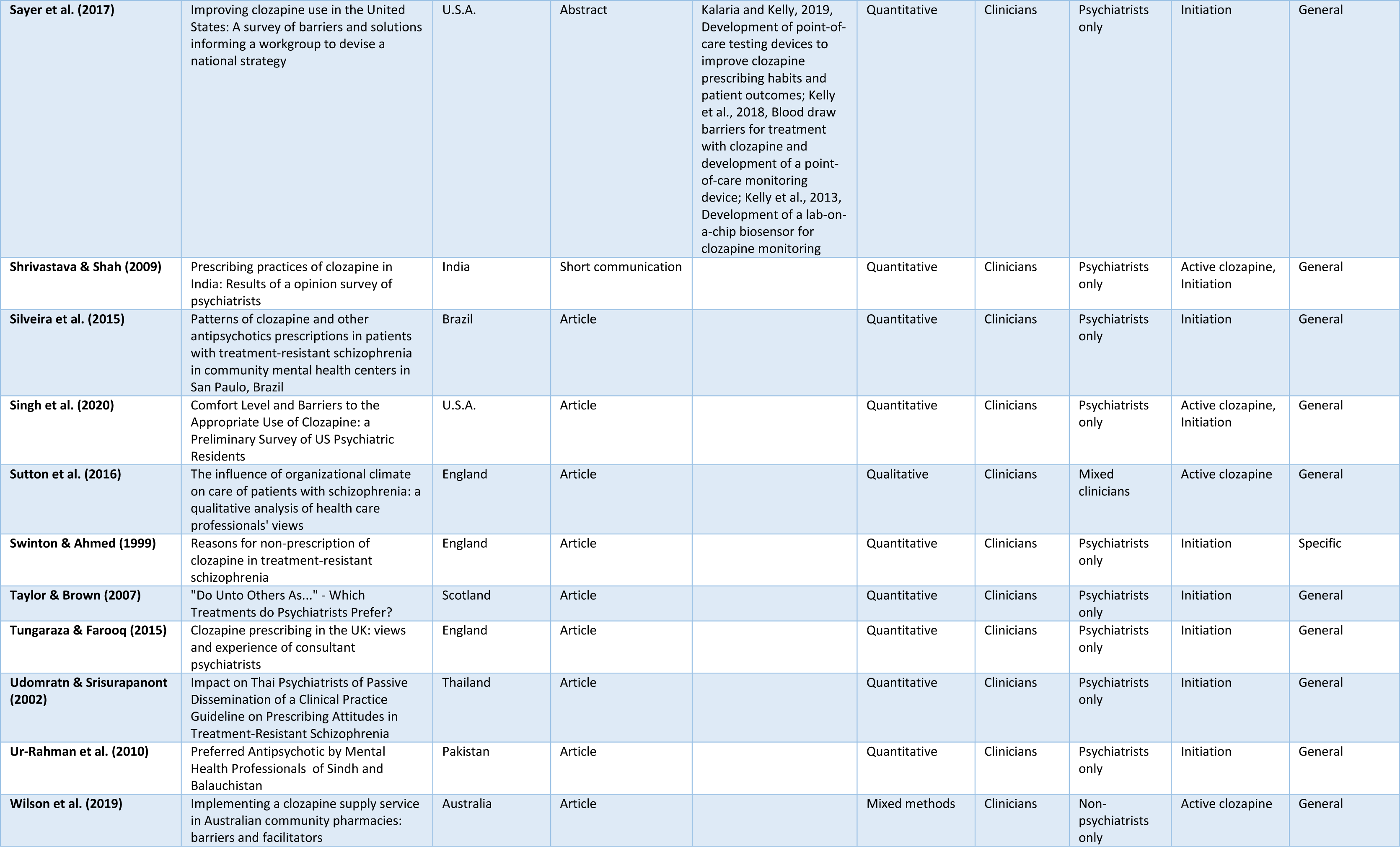
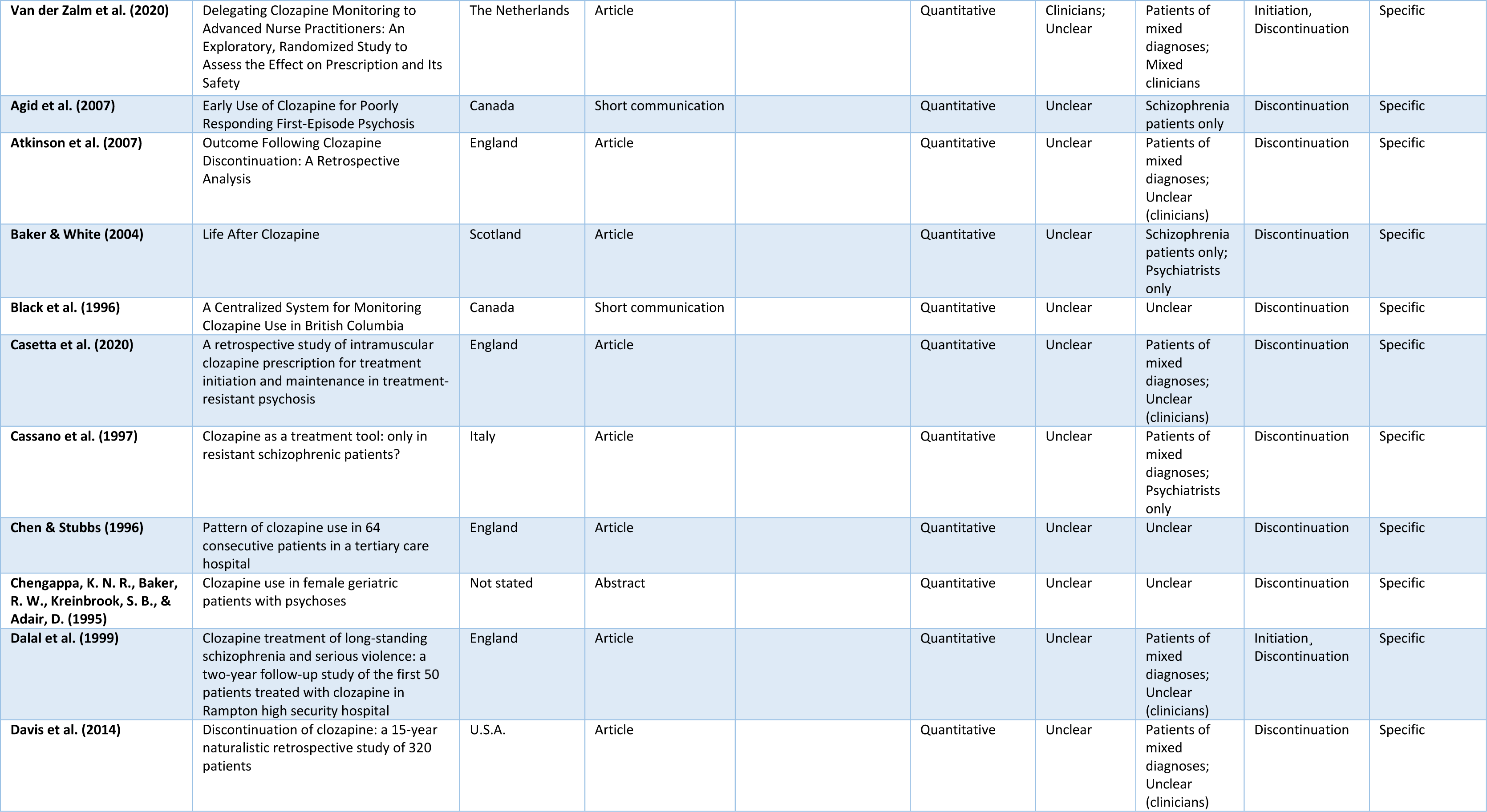
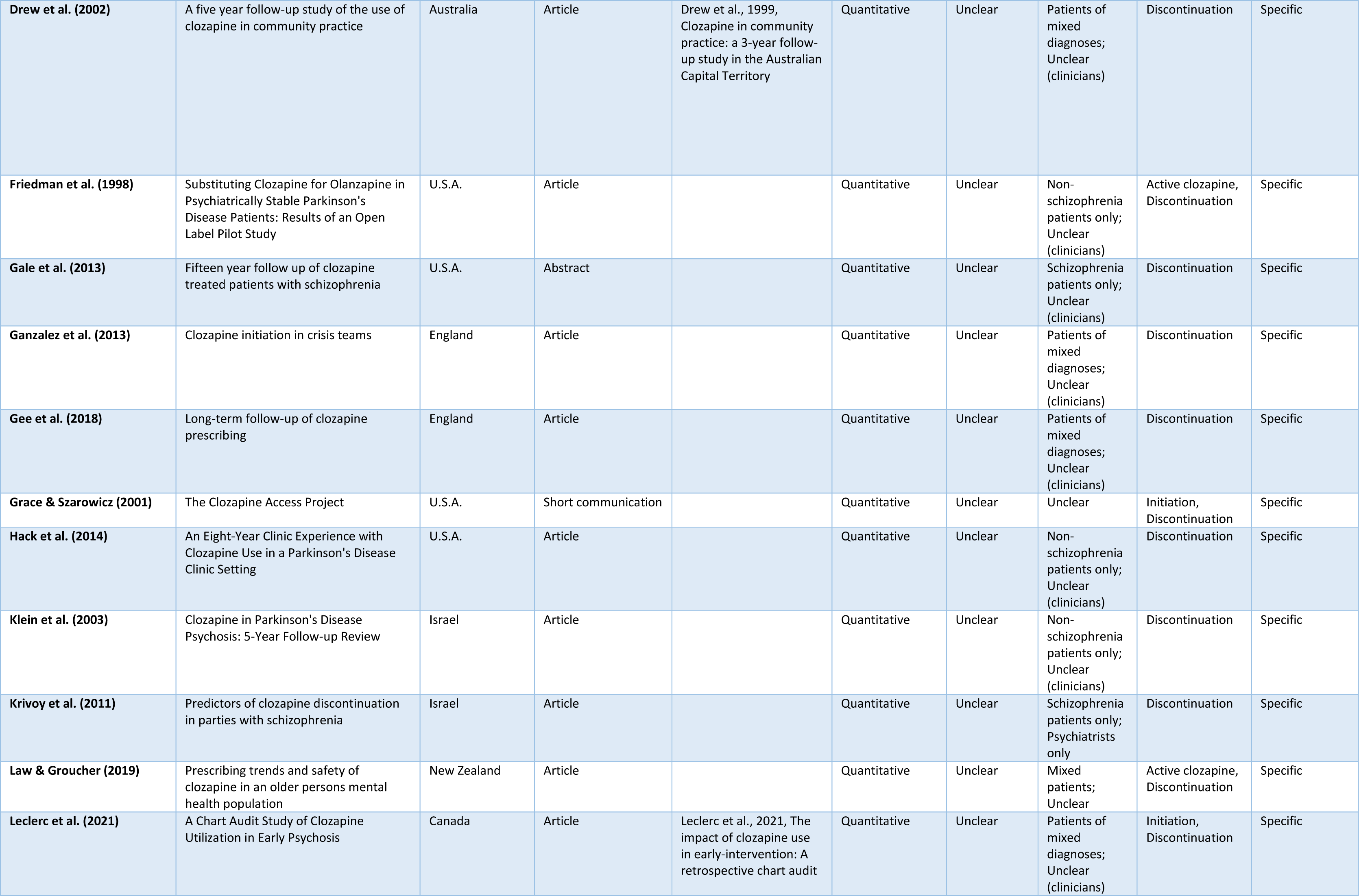
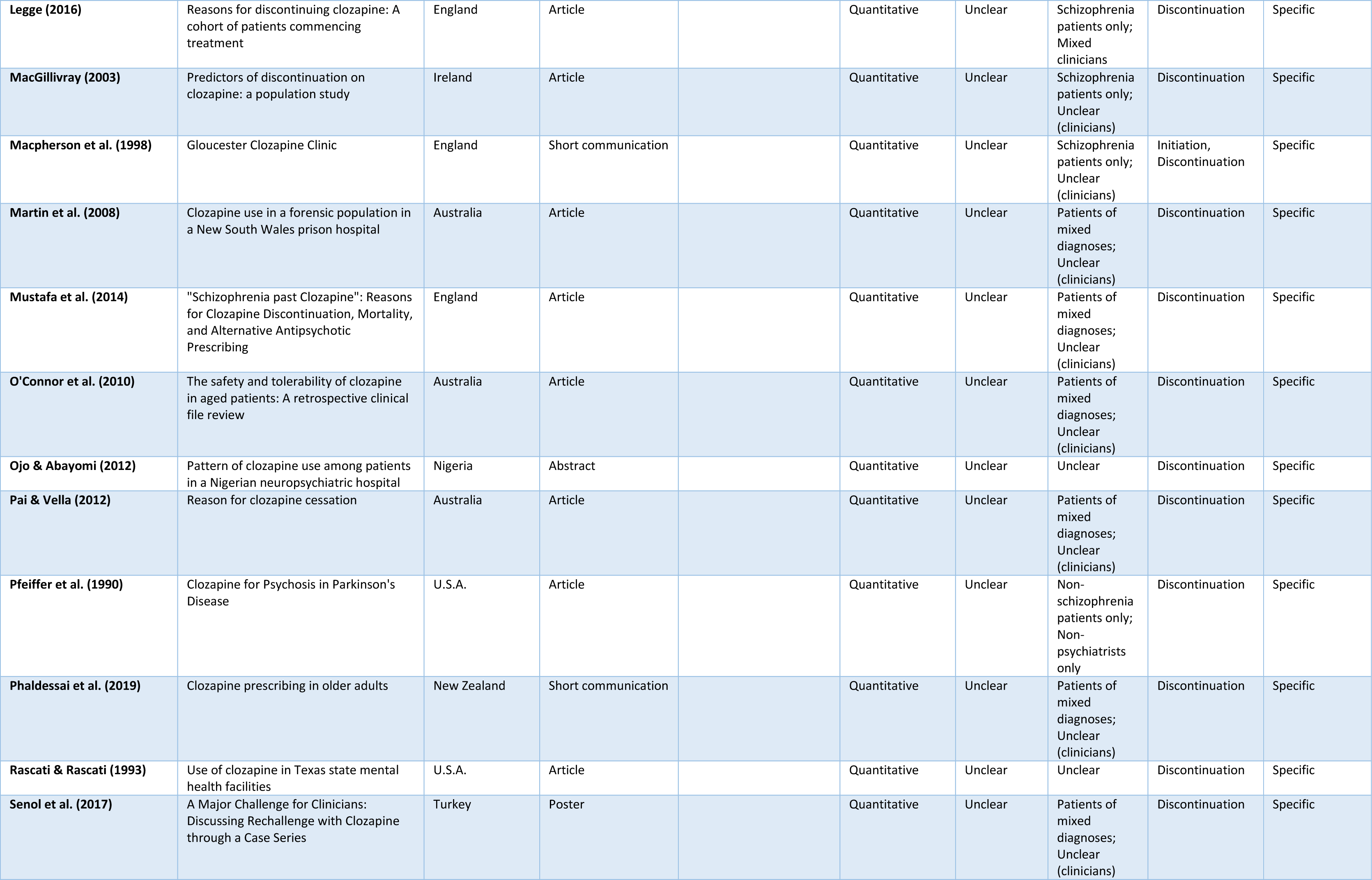
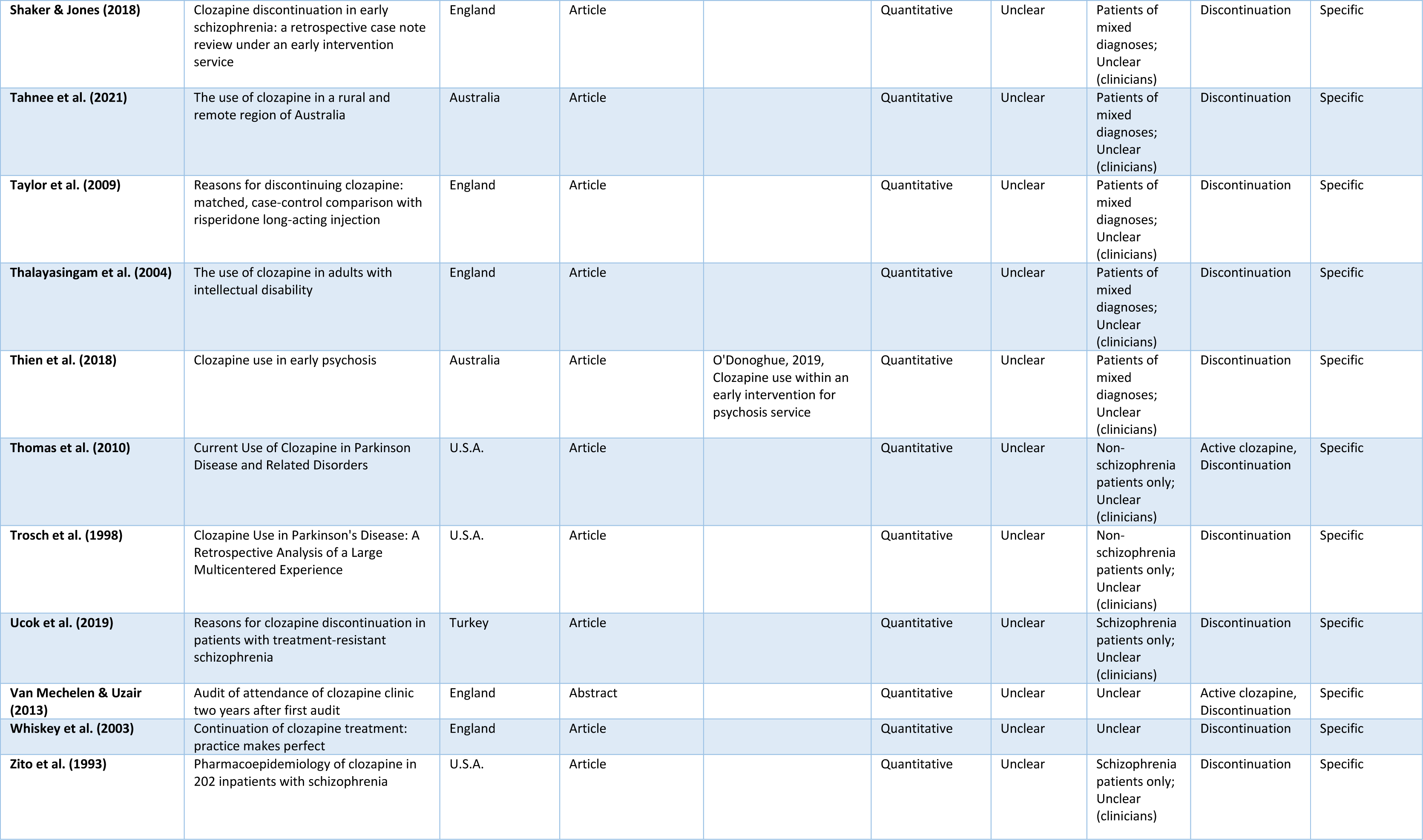
Included studies.

The search identified 146 studies^22–25 29–39 41–43 56–183^, represented by 158 sources of evidence^22–25 29–39 41–43 56–195^ and 57 countries across six continents. Of these, 115 studies were extracted as articles, two as reports, 14 as short communications, one as an article summary, 13 as abstracts, and one as a conference poster. The year of publication was between 1990 and 2021. One-hundred-and-thirty-eight studies used a quantitative design (i.e. survey questionnaires, rating scales, case-note reviews, etc.), 13 used a qualitative design (interviews), one study used both, and six studies used a mixed-methods design. Individual, in-depth study characteristics are provided in the Supplementary data file 2 (S2, “Individual study data”, doi.org/10.17605/OSF.IO/HKBSG) retrievable from the Open Science Framework repository ^196^.

Thirty-nine of the studies^22–25 29–39 41 43 44 71 86 99 100 102 105 109 110 117 119 122 130–132 134 136 137 182 188 197 198^ have been included in previous systematic reviews^17–21 27 28^ addressing patients’ and/or clinicians’ perspectives on clozapine treatment.

### 3.3. Distribution of studies according to the origin and content categories of explored perspectives

Figure 2 provides a graphic presentation of the distribution of studies.

**Figure 2.**
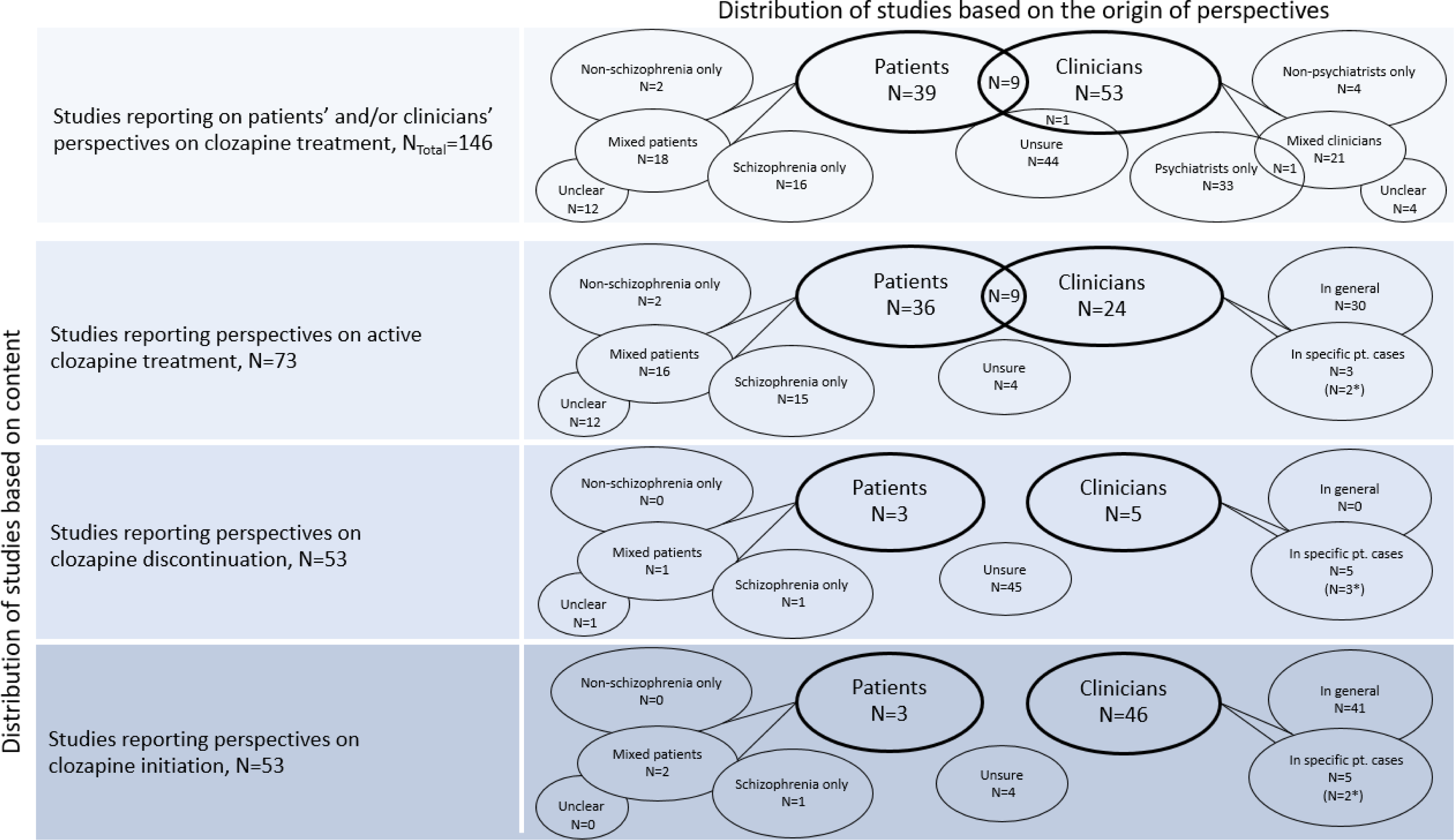
Graphic presentation of the scope of studies reporting on patients’ and/or clinicians’ perspectives on clozapine treatment. Notes The sum of studies depicted in row 2-4 does not correspond the total number (N_Total_) of included studies (row 1). Some studies report perspectives on more than one content category (e.g. on both active treatment and initiation), i.e. the same study may be counted multiple times, representing different content categories (rows). Some studies report on perspectives from more than one study population (e.g. both patient and clinician perspectives). The number of studies reporting “overlapping” perspectives from different populations is shown in the intersection between two ovals. In several studies, the reported perspectives could not be assigned to either patients or clinicians, e.g. due to the indirect/second hand origin of data from case files. The origin of perspectives in these studies are labelled “Unsure”. * The number of clinician studies reporting case-specific psychiatrist perspectives.

Of the 146 included studies, 39 reported on patient perspectives^22 24 29–38 41–43 56–79^, nine reported on both patient and clinician perspectives^23 25 39 80–85^, and 54 reported on clinician perspectives ^86–139^. In 45 studies ^139–183^, the origin of some or all reported perspectives was unclear (Figure 2, row 1).

Of the total 63 studies reporting on clinician perspectives, 34 reported on perspectives belonging to psychiatrists^86–93 95–97 102 103 106 107 109 113 116 119 122–132 134–137^ four on perspectives belonging to non-psychiatrists only^39 94 98 138^ and 22 on perspectives belonging to a mixture of different clinicians^25 80 82 84 85 88 99–101 104 105 108 110–112 114 115 117 118 121 133 139^. In four studies^23 81 83 120^, the type of clinicians was unclear (Figure 2, row 1).

Of the total 48 studies on patient perspectives, 16 reported on perspectives belonging to schizophrenia patients in specific^29 30 35 37 39 43 58 63 65 70 71 75–78 84^, two on non-schizophrenia patients only^67 74^, 18 on patients of mixed diagnoses^23–25 34 36 38 41 42 57 60–62 68 69 72 73 79 82^ and in 12 studies^22 31–33 56 59 64 66 80 81 85 193^ the type of patients was unclear (Figure 2, row 1).

Seventy-three studies reported on perspectives related to active/ongoing clozapine treatment^22–25 29–39 42 43 56–76 78 80–85 89–96 98 100 102–104 108 110 115 117 119 122 126 130 132 133 138 151 159 178 181^ (Figure 2, row 2).

Forty-five studies reported on clozapine patients’ attitudes towards their treatment^22–25 29–39 42 43 56–76 78 80–85^, of which 15 studies reported specifically upon experiences belonging to schizophrenia patients^29 30 35 37 39 43 58 63 65 70 71 75 76 78 84^. Thirty-three studies reported upon clinicians’ perspectives on clozapine treatment^23 25 39 80–85 89–96 98 100 102–104 108 110 115 117 119 122 126 130 132 133 138^, of which three reported upon clinicians’ perspectives on clozapine treatment in relation to specific patient cases^39 90 103^. In four studies ^151 159 178 181^, the origin of perspectives on active treatment was unclear.

Fifty-three studies reported upon perspectives on clozapine discontinuation^33 78 79 90 116 118 120 125 139–183^ (Figure 2, row 3). Three studies reported upon patients’ reasons for discontinuing clozapine treatment^33 78 79^, and five studies reported upon clinicians’ perceived reasons for clozapine discontinuation^90 116 118 120 125^, all of them in specific patient cases. Forty-five studies reported upon reasons for discontinuation but could not be assigned to either patient or clinician perspectives due to the case-note origin of the data^139–183^.

Fifty-three studies reported upon perspectives on clozapine initiation^41 77 190 82 86–93 95–102 104–107 109–114 117 118 120–124 126–132 134–137 139 148 155 160 163^ (Figure 2, row 4). Three studies reported upon patients’ attitudes towards clozapine commencement^41 77 190^, whereas 46 studies reported upon clinicians’ perspectives on clozapine initiation^82 86–93 95-102 104-107 109-114 117 118 120-124 126-132 134-137 139^. In five of these studies^90 118 120 134 139^, the clinician perspectives were related to specific cases of clozapine initiation. In four studies^148 155 160 163^, the origin of perspectives was unclear.

### 3.4. Characteristics of explored patient perspectives

In the 45 studies on clozapine patients’ attitudes towards their medication^22–25 29–39 42 43 56–76 78 80–85^, 10 different sub-categories of perspectives were explored (Figure 3a.), the most frequent ones being the Perceived burden of treatment (n=36) and Perceived efficacy of treatment (n=34).

**Figure 3.a.**
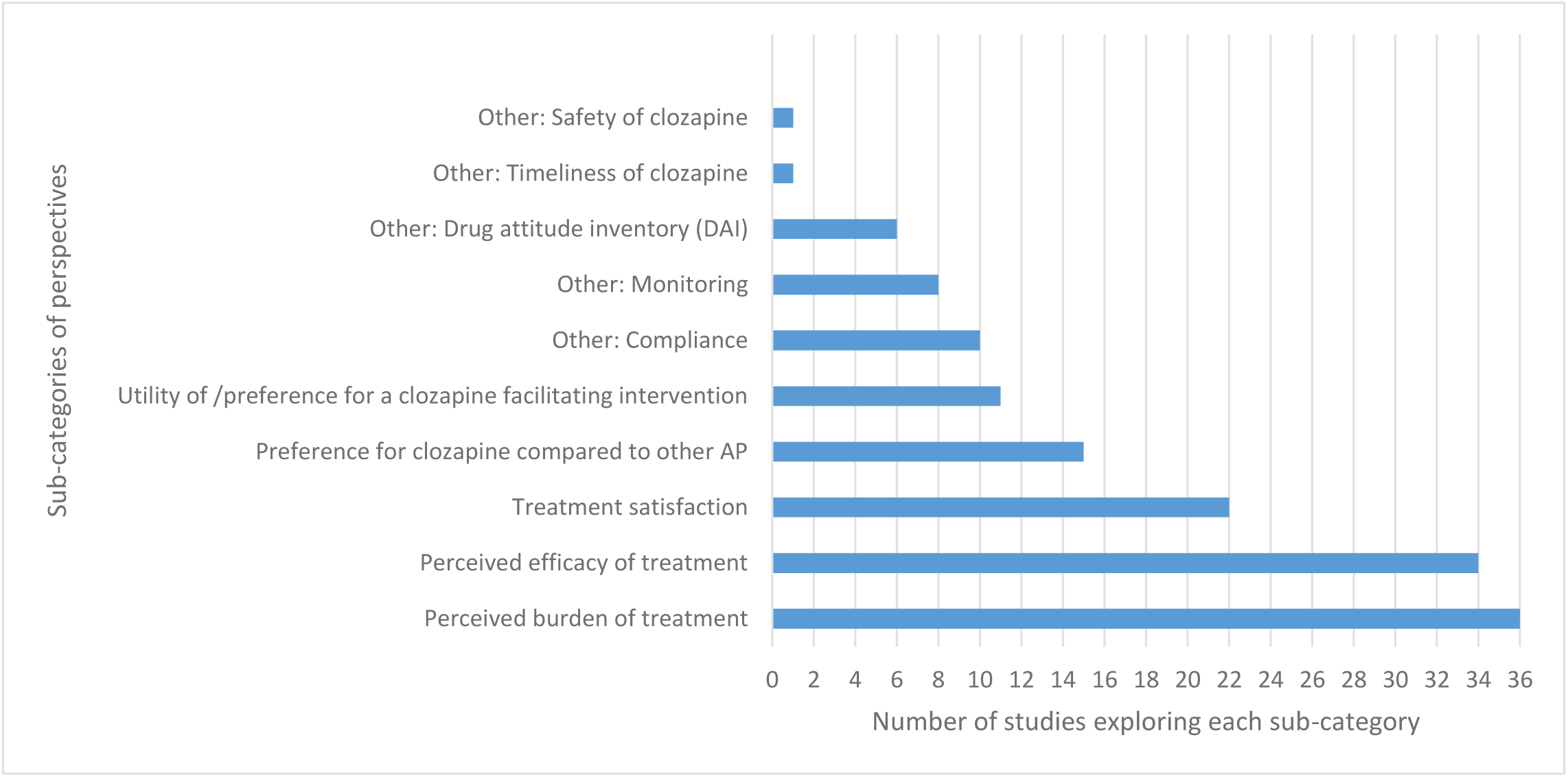
The distribution of explored sub-categories of perspectives in studies on patient perspectives on active/ongoing clozapine treatment (N= 45).

The patients’ perspectives on clozapine treatment were overall positive, with the majority of patients expressing high efficacy- and satisfaction ratings with clozapine and a preference for clozapine compared to their previous treatment - regardless of diagnosis. Adverse side effects and the need for blood monitoring was generally accepted amongst all groups of patients due to the perceived efficacy of clozapine treatment. However, patients tended to prefer Point-Of-Care (POC) devices for finger prick blood sampling to conventional venipuncture - as well as other interventions aiming at simplifying treatment (the categories of assessed interventions are shown in Figure 3.b.). Patients with organic psychosis/neurological conditions seemed more vulnerable to adverse side effects and to the logistic barriers to monitoring (See the Supplementary data file (S2)^196^, under “Key findings”, for individual, in-depth study findings).

**Figure 3.b.**
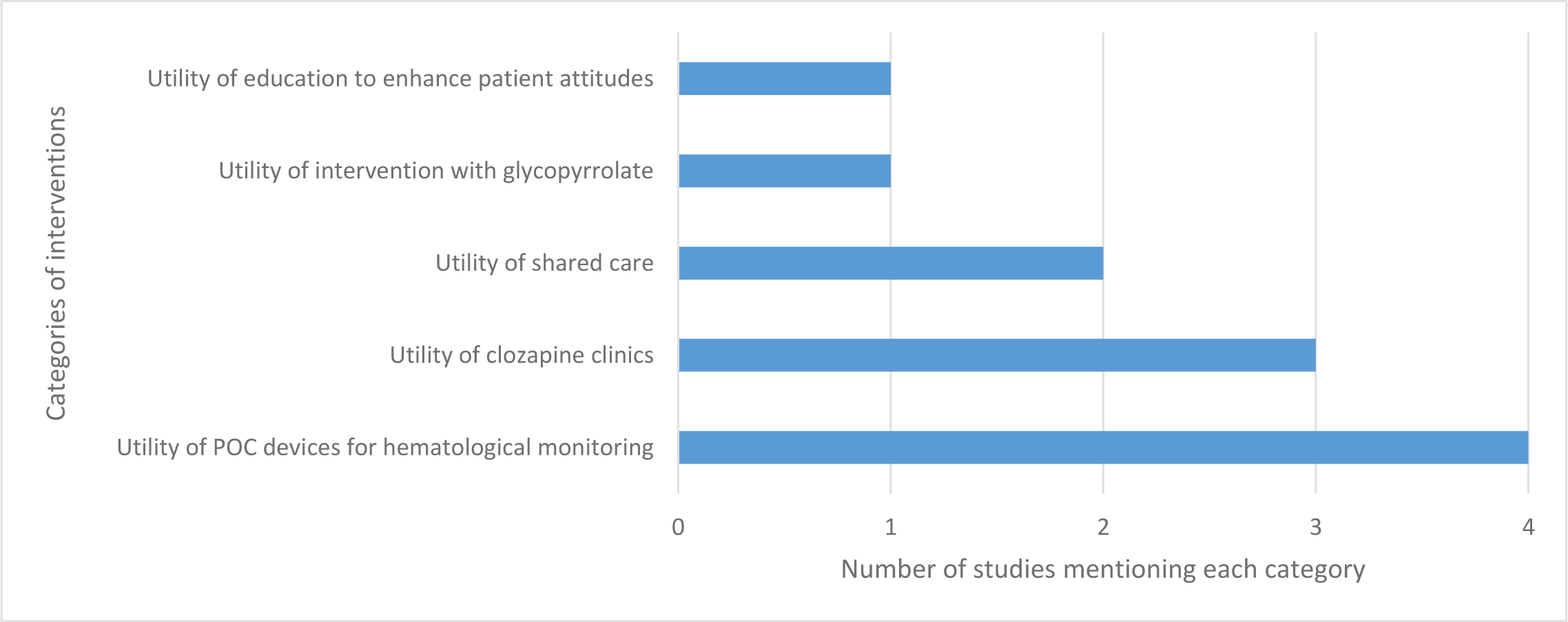
The distribution of mentioned clozapine-facilitating interventions in studies on patients’ perspectives on active/ongoing clozapine treatment (N=11).

In the three studies reporting on patients’ perspectives on clozapine discontinuation^33 78 79^, the reasons for discontinuation were explored. Five categories of reasons were mentioned (Figure 4), with Adverse side effects (n=3) and Lack of efficacy (n=2) being the most frequent ones. Lack of efficacy was the most prominent reason in two out of three studies^78 79^ (S2^196^, Key findings).

**Figure 4.**
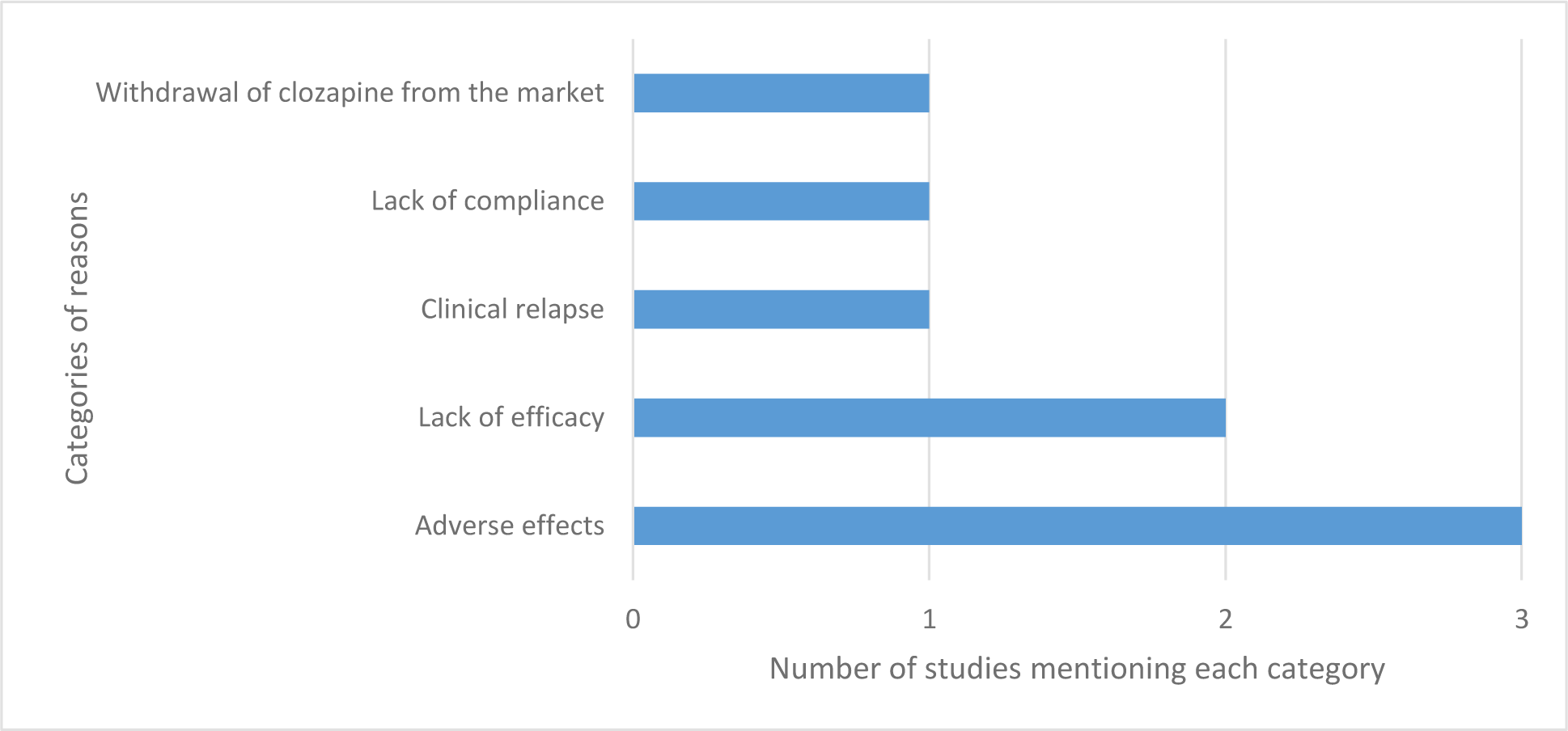
Distribution of categories of reasons for clozapine discontinuation mentioned in studies on patient perspectives on clozapine discontinuation (N=3).

In the three studies on patients’ perspectives on clozapine commencement^41 77 79^, six sub-categories of perspectives were explored (Figure 5). The sub-category Patients’ reasons for refusal was, as the only sub-category, assessed in two studies^41 77^; however, only fully reported upon in one of them – a study on acutely unwell inpatients with schizophrenia or schizoaffective disorder^41^. The prospect of admission as a requirement for clozapine commencement seemed to be the main barrier to acceptance for most patients in this study. Blood monitoring was not the most prominent reason for refusal in any of the two studies (S2^196^, Key findings).

**Figure 5.**
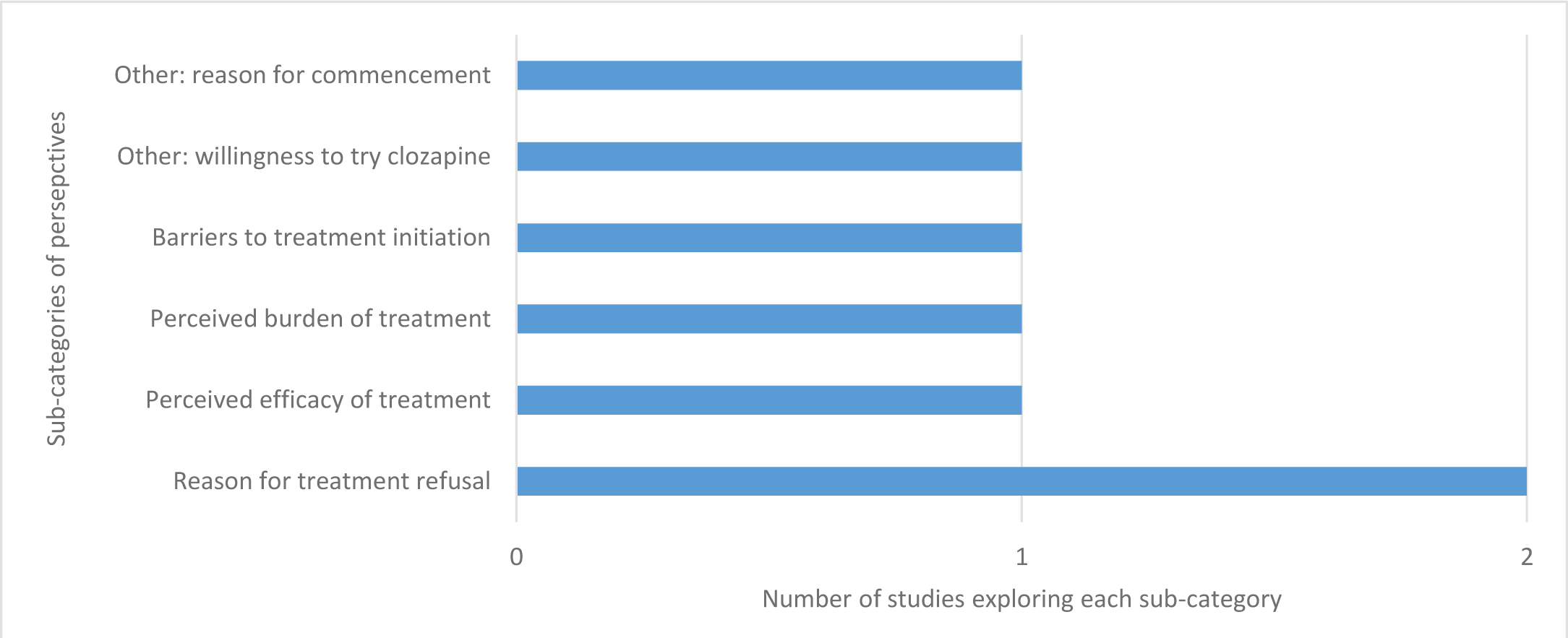
Distribution of explored sub-categories of perspectives in studies on patient perspectives on clozapine commencement (N=3).

### 3.5. Characteristics of explored clinician perspectives

In the 33 studies on clinician perspectives on active clozapine treatment^23 25 39 80–85 89–96 98 100 102–104 108 110 115 117 119 122 126 130 132 133 138^, 13 different sub-categories of perspectives were explored (Figure 6.a.). The most frequently explored sub-categories of perspectives were Perceived efficacy of treatment (n=18) and Utility of /preference for a clozapine facilitating intervention (n=16). In the latter, eight different categories of interventions aiming at improving treatment utility, satisfaction or adherence were assessed (Figure 6.b).

**Figure 6.a.**
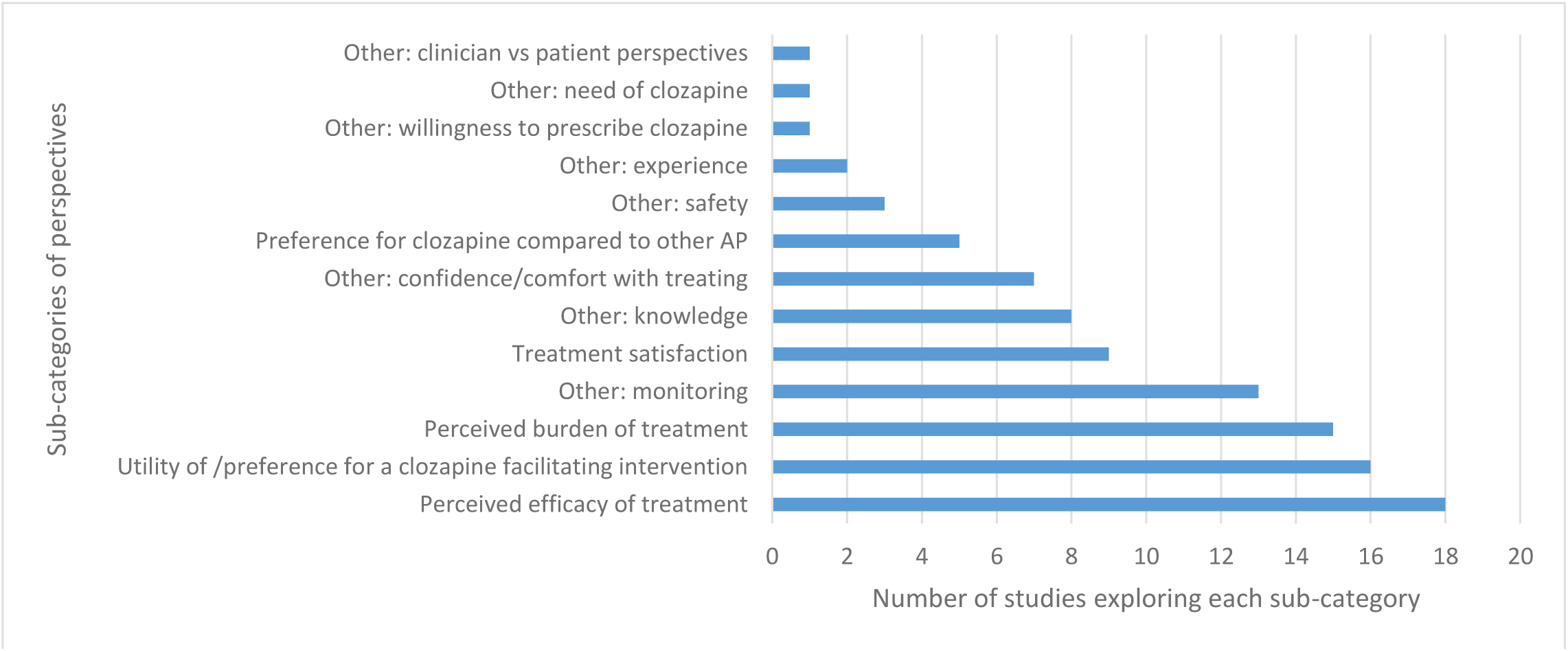
Distribution of explored sub-categories of perspectives in studies on clinician perspectives on active/ongoing clozapine treatment (N=33).

**Figure 6.b.**
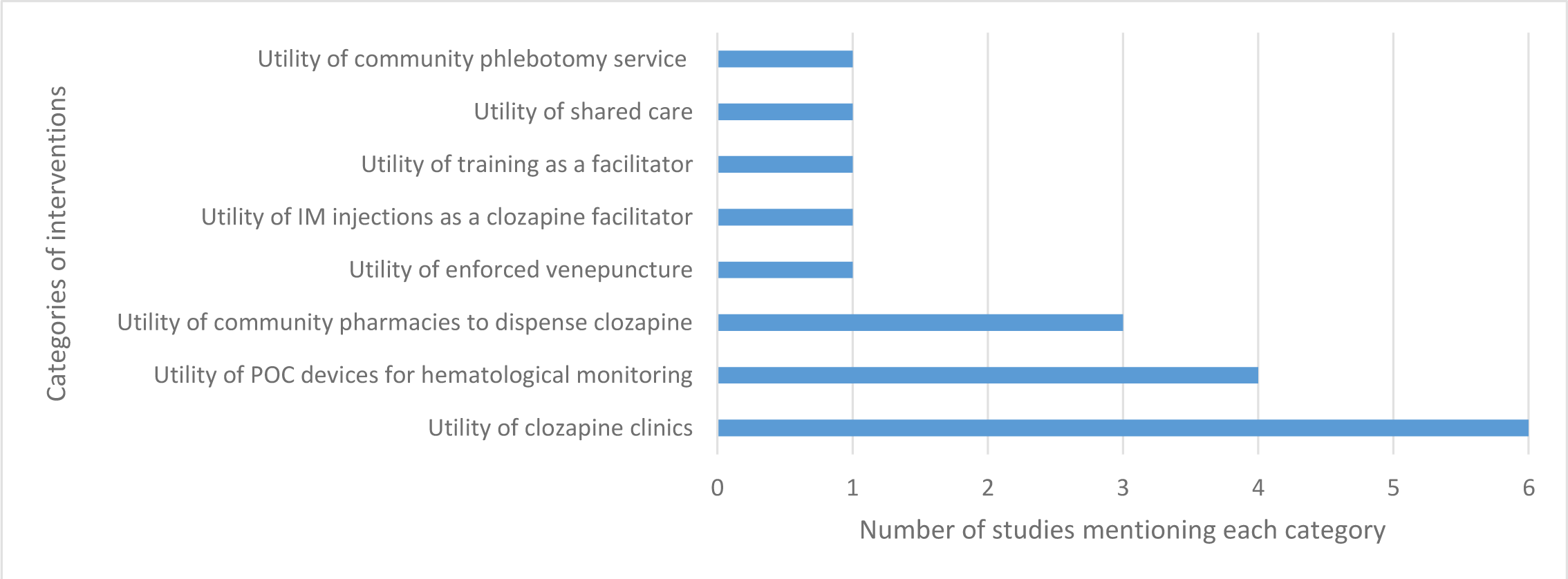
Distribution of mentioned clozapine-facilitating interventions in studies on clinicians’ perspectives on active/ ongoing clozapine treatment (N=16).

Most studies on clinician perspectives in relation to active clozapine treatment were studies of general perspectives (n=30); however, three studies^39 90 103^ reported clinician perspectives in relation to specific patient-cases of clozapine treatment. The sub-categories of explored perspectives in these studies were Efficacy of treatment (n=3) and Perceived burden of treatment (n=1).

Overall, the clinicians perceived clozapine to be efficient, but of great burden to the patients in terms of adverse side effects and blood monitoring requirements. The clinicians were consistently in favor of POC devices for capillary hematological monitoring over conventional venous sampling as this was considered to increase both patient and clinician satisfaction with treatment. Other types of facilitating interventions were also considered relevant although assessed less frequently or, as in the case of clozapine clinics, with mixed enthusiasm (S2^196^, Key findings).

In the five studies on clinician perspectives on clozapine discontinuation^90 116 118 120 125^, the reasons for clozapine discontinuation were assessed. In total, nine different categories of reasons were mentioned (Figure 7), of which No/ poor response, Patient’s refusal to take clozapine and/or do bloodwork, Non-compliance with clozapine and Adverse side effects were the most frequently mentioned ones, each in four studies. However, Adverse side effects was the most prominent reason in most studies (n=3) and non-compliance in two studies. See also S2^196^, Key findings, for more information on individual study findings.

**Figure 7.**
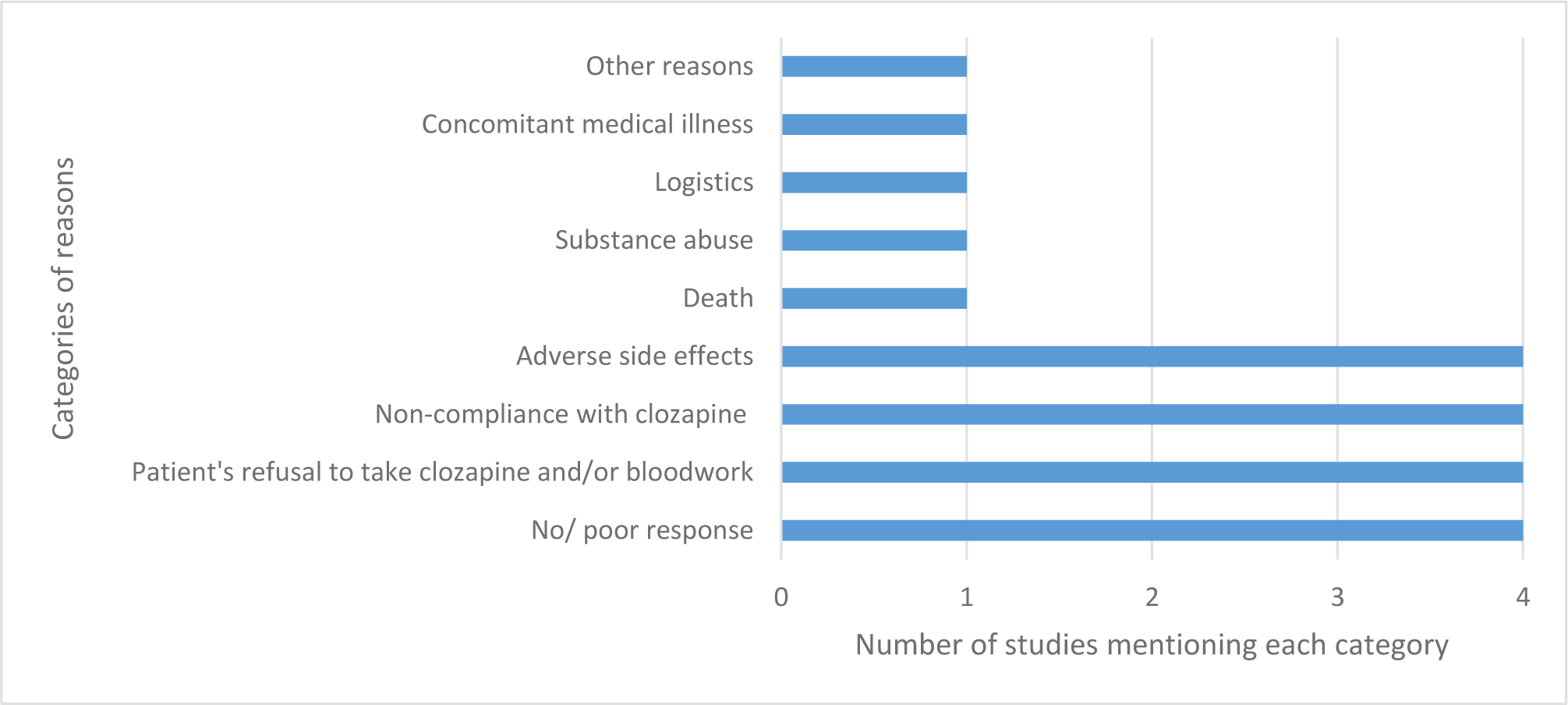
Distribution of categories of reasons for discontinuation mentioned in studies on clinician perspectives on clozapine discontinuation (N=5).

Forty-six studies explored clinician perspectives on clozapine initiation^82 86–93 95–102 104–107 109–114 117 118 120–124 126–132 134–137 139^, most of them (n=41) in general terms. Thirteen sub-categories of perspectives were explored (Figure 8.a.), the most frequent ones being Barriers to treatment initiation (n=29) and Utility of /preference for a clozapine facilitating intervention (n=23). Eighteen categories of barriers to initiation were mentioned (Figure 8.b.), of which Concerns about adverse side effects was the most frequently mentioned one (n=20). Seventeen sub-categories of interventions were mentioned as facilitators of clozapine initiation (Figure 8.c.), most of them referring to more training in clozapine treatment (n=6) and the outsourcing of clozapine initiation (n=6), monitoring and/or prescribing (n=6).

**Figure 8.a.**
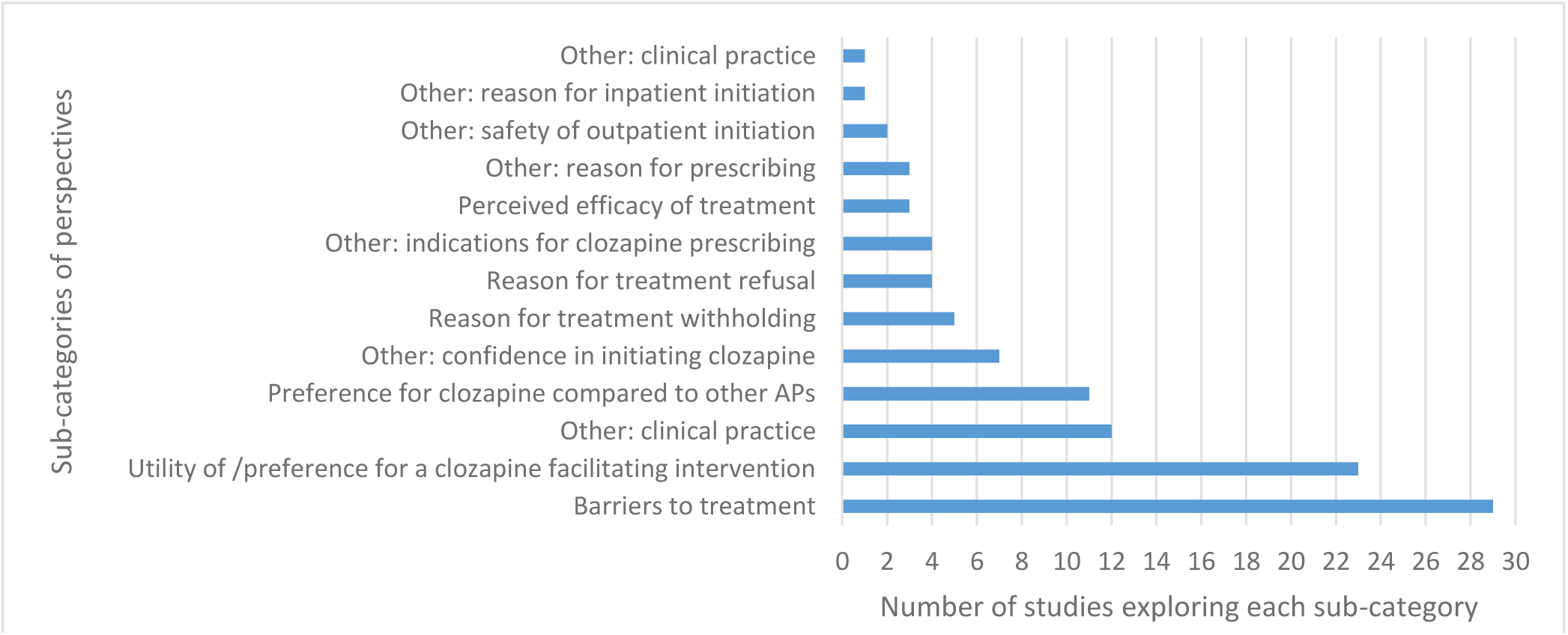
Distribution of explored sub-categories of perspectives in studies on clinicians’ perspectives on clozapine initiation (N=46).

**Figure 8.b.**
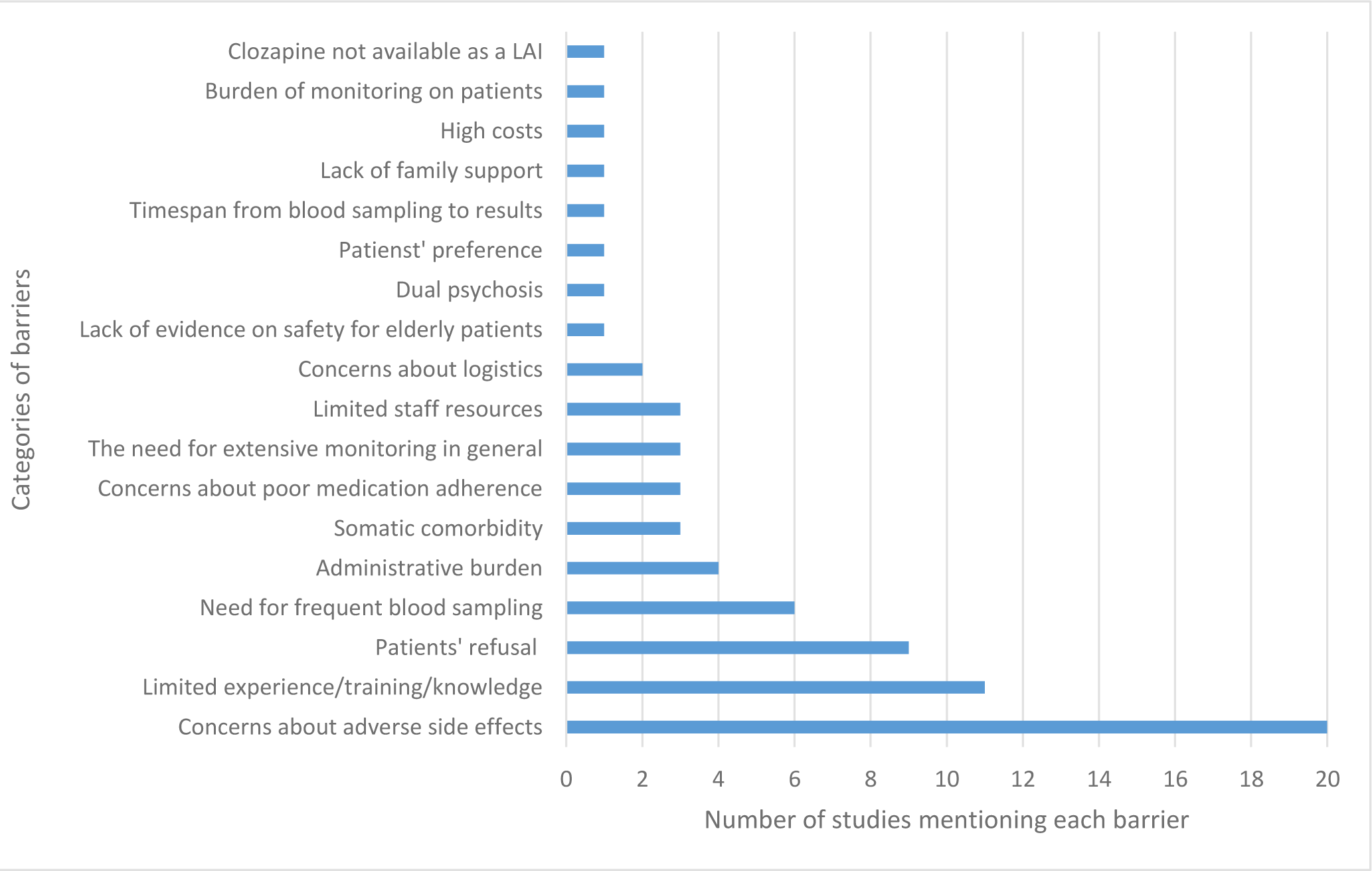
Distribution of mentioned barriers

**Figure 8.c.**
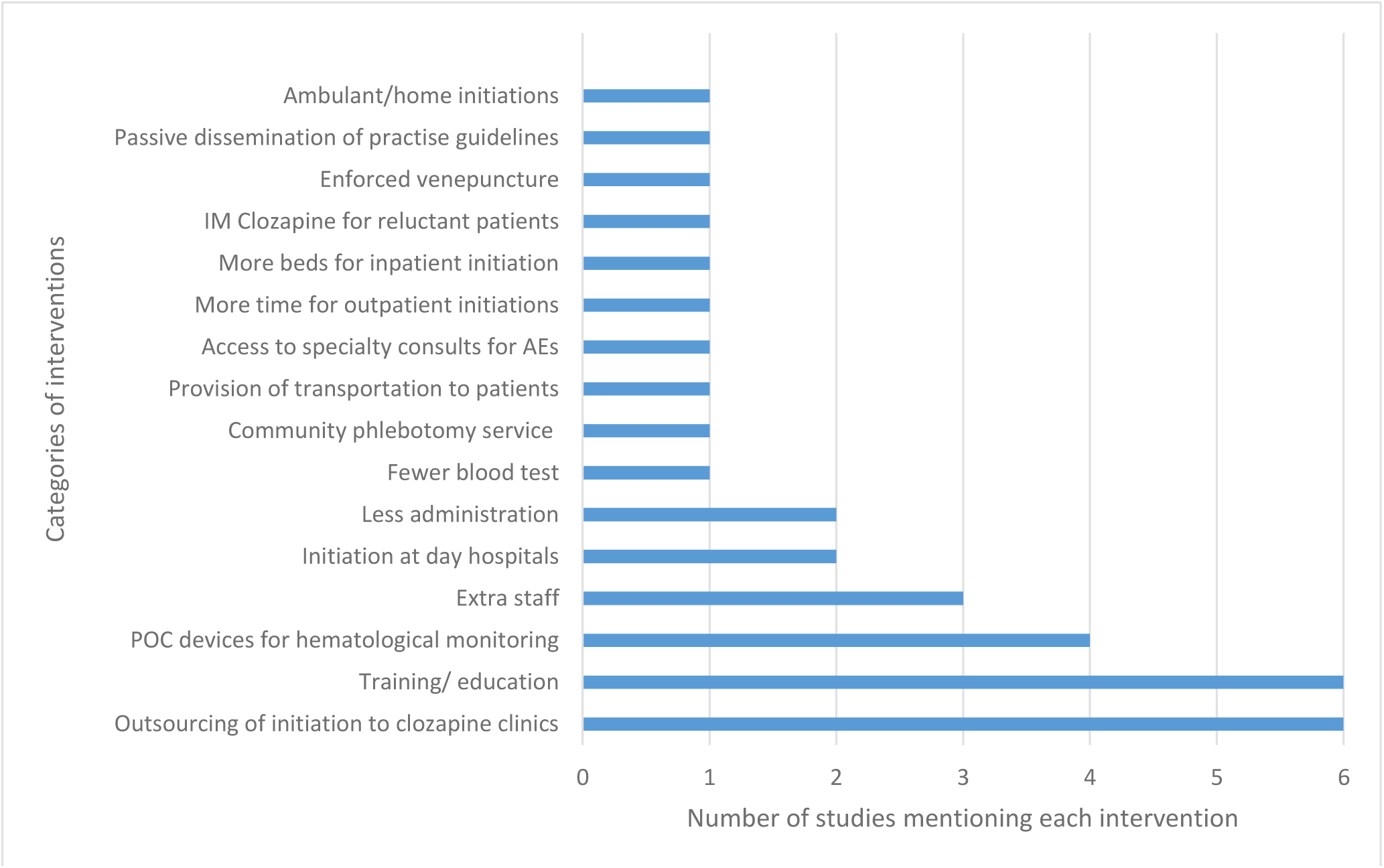
Distribution of interventions mentioned as facilitators of clozapine initiation in studies on clinicians’ perspectives on clozapine initiation (N=23).

Five studies explored clinician perspectives on clozapine initiation in relation to specific patient cases^90 118 120 134 139^, two of them reported perspectives belonging to psychiatrists in specific^90 134^. The sub-categories of explored perspectives in the case-specific studies were Reasons for treatment withholding (n=4), Utility of /preference for a clozapine facilitating intervention (n=2), Barriers to initiation (n=2), Other: Reason for inpatient initiation (n=1), Other: Indication for treatment (n=1) and Other: Clinical practice (n=1).

Two of the case-specific studies relied on case note data^120 139^, whilst three studies used a more direct approach of assessment^90 118 134^. In two of these “direct” studies^118 134^, the patients were inpatients and in one study^90^, the patients’ status as in- or outpatients was not stated.

In the four case-specific studies exploring Reasons for treatment withholding^118 120 134 139^, six categories of reasons were mentioned (Figure 8.d.). The most frequently mentioned ones were Expected non-compliance with either drug or blood monitoring (n=2), Risks outweighs benefits (n=2) and Somatic issues (n=2).

**Figure. 8.d.**
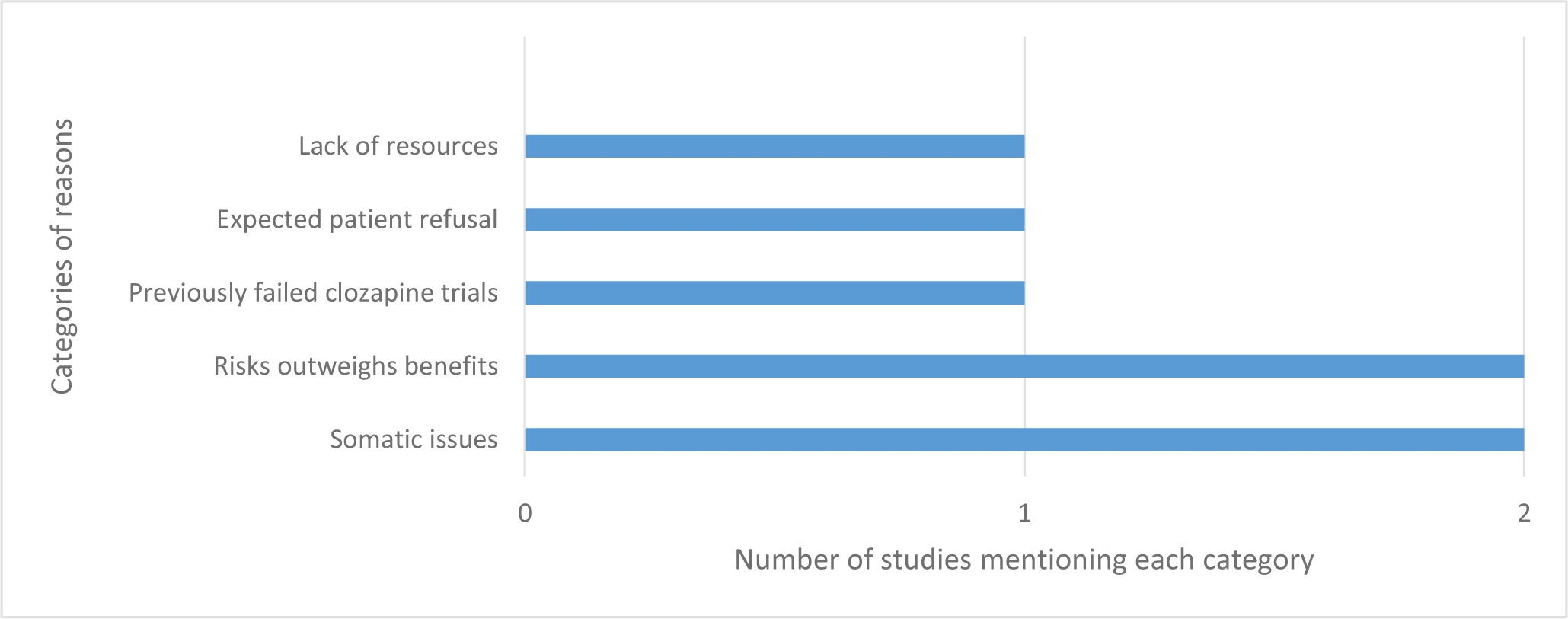
Distribution of reasons for treatment withholding mentioned in studies on clinicians’ perspectives on clozapine initiation (N=5).

Data on psychiatrists’ reasons for clozapine withholding in specific cases was only reported upon in one study of inpatients^134^, in which expected patient refusal or non-compliance with either drug or blood monitoring were the most frequently stated reasons.

## 4. Discussion

### 4.1. Summary of evidence

With this scoping review, we aimed to map the scope of primary studies reporting upon patients’ and/or clinicians’ perspectives on clozapine treatment. The map was intended to provide an overview of the types of perspectives that have been explored as well as to identify any cluster formations and/or gaps in the existing research. We included 146 studies, of which only 39 had been included in previous systematic reviews addressing patients’ and/or clinicians’ perspectives on clozapine treatment. We showed that the research in this area tends to repeat itself; most studies on the subject could be assigned to clinician perspectives (n=63, 43%) and most of these studies centered around general perspectives on clozapine initiation (n=42, 67%). Forty-eight studies (33%) reported upon patients’ perspectives, the majority of them (n=45, 94%) on clozapine patients’ attitudes towards their ongoing treatment. In contrast, only a few studies reported on patients’ perspectives on clozapine commencement (n=3, 2%) or clinicians’ perceived reasons for non-clozapine treatment in specific patient cases (n=5, 3%).

In line with established scoping review conventions ^45–49^, no formal critical appraisal has been conducted regarding the sources of evidence. However, several factors affected the analysis of scope and clusters. Of the five case-specific studies on clinician-perceived reasons for non-clozapine treatment, only three studies used a direct approach to the assessment of reasons, the others relied on case-note data. Moreover, only two of these studies reported specifically on prescriber-perceived reasons and none of the studies could elucidate the reasons for clozapine withholding for clozapine-eligible outpatients – only inpatients.

Of the three studies on patients’ attitudes towards clozapine commencement, only one study assessed the patients’ willingness to try clozapine and the factors affecting their choice. As with the studies on clinicians’ reasons for withholding, this was a study on inpatients.

Furthermore, a substantial number of studies reported upon reasons for clozapine discontinuation (n=53, 36%), however, due to the second- or even third-hand origin of data from case notes, the reported reasons could not be assigned to either patients or clinicians in most studies (n=45, 85%). In some studies, the main reasons were divided into patient and clinician categories, however, interpreted as such by the authors themselves. Consequently, only a few studies reflected the direct assessment of patients’ or clinicians’ perspectives on clozapine discontinuations.

As this review underpins, the underutilization of clozapine has been a concern for decades and a vast number of studies have sought to understand both the patients’ and the clinicians’ experiences or perceptions of clozapine treatment, the clinicians’ perceived reasons for barriers or facilitators of its usage as well as the reasons leading to clozapine discontinuation. Despite that, clozapine remains both underutilized ^12^ and initiated too late in the treatment course with major consequences to the patients’ long-term prognosis^199^. One explanation for the lack of change could be the repetitive pattern of previous research described above. We reckon that evidence on general clinician perspectives on barriers to clozapine prescribing has met its saturation point and that both patient and clinician perspectives on active clozapine treatment are well documented by now. Additional studies in these areas will merely repeat what is already known - although still highly relevant to assess in a clinical context. However, there is an apparent gap of evidence on clozapine-naïve patients’ attitudes towards clozapine commencement, in particular in regards to eligible outpatients, as well as of direct (i.e. not case-note derived) assessments of clinicians’ reasons for clozapine withholding in specific cases of eligible outpatients. These kinds of studies could provide the information needed for a successful increase in clozapine utilization.

Furthermore, due to the limited number of studies and categories of perspectives assessed in terms of patients’ and clinicians’ perspectives on clozapine discontinuation, future studies, exploring both the patients’ and the clinicians’ needs in terms of clozapine discontinuation vs. continuation or re-challenge in specific patient-cases, could provide valuable insights for treatment optimization.

Since the beginning of this work, we have conducted our own study on case-specific clinician reasons for clozapine withholding^200^ to address one of the identified gaps in research. A renewed Google Scholar search conducted on January 3^rd^, 2024, further revealed the publication of two new studies on patient perspectives towards clozapine discontinuation^201 202^. No other published studies within the identified under-prioritized areas were found. Instead, eight additional studies on general clinician perspectives on clozapine treatment^203–210^, three additional studies on clozapine-patients’ (and their clinicians’) attitudes towards their ongoing treatment^40 211 212^, and three additional studies on case-note derived reasons for clozapine cessation or withholding^213–215^ appeared, ratifying the repetitive behavior of current research and the importance of disseminating these results in order to direct future research towards more meaningful and less well-described areas.

### 4.2. Strength and limitations

Strengths: this scoping review has been developed in accordance with established scoping review methodology to ensure methodological rigor. The prospective registration and publication of the review protocol has ensured transparency of the review process. The three-stepped search strategy, involving a broad range of search terms and relevant databases and sources of grey literature, has ensured a comprehensive search of the literature and the multiple booster searches on Google Scholar have ensured a continued (and recent) update on the scope of literature. Limitations: The original database search was conducted in June 2021. Furthermore, the comprehensive search strategy, including terms as Perspective, Attitude and Subjective, entailed the risk of identifying an overwhelmingly large amount of studies, including commentaries and letters expressing individual opinions. As a means to ensure both feasibility and a certain level of scientific evidence, the exclusion of such data sources was deemed necessary. Due to feasibility reasons, the search was restricted to publications in English. However, studies providing résumés, abstracts or posters in the English language were included, even if the full texts were retrievable only in the native language. Still, these pragmatic decisions may have precluded the identification of some relevant insights and studies.

### 4.3. Conclusions

A substantial number of studies reporting on patients’ and/or clinicians’ perspectives on clozapine treatment were identified, most studies without previous inclusion in systematic reviews. The mapping of studies revealed a repetitive form of current research with a cluster formation surrounding clozapine patients’ attitudes towards their ongoing treatment, general clinician perspectives on active clozapine treatment, and general clinician perspectives on barriers to clozapine initiation. However, three apparent gaps in research were identified: 1) clozapine eligible, yet clozapine-naïve, outpatients’ attitudes towards clozapine commencement, 2) assessments of clinicians’ reasons for clozapine withholding and perceived facilitators of clozapine treatment in specific patient-cases, and 3) direct (i.e. not case-note derived) assessments of both patient and clinician perspectives on clozapine discontinuation, continuation and re-challenge in specific patient-cases. Future studies within these areas of research could provide the evidence needed to turn clozapine from underutilization.

## Declarations

### Ethics

The scoping review does not require ethics approval.

### Patient and public involvement

This review was developed without public or patient involvement.

## Acknowledgments

We thank our research librarian, Trine Kæstel, Psychiatric Research Unit, Region Zealand Psychiatry, Denmark, for assisting with the search strategy.

## Contributors

Authors MIJ, OJS, SFA, JN and ES participated in conceptualizing the review. Author MIJ developed the search strategy in consultancy with a research librarian (see “Acknowledgements”), wrote the protocol, performed the search and managed the data. Authors JPS and MIJ performed the screening and data extraction in collaboration; author OJS acted as a third-party consultant in cases of doubt. Author MIJ wrote the first draft of the manuscript. All authors have critically reviewed the manuscript and approved the final version for submission.

## Data availability statement

The data that support the findings of this study are retrievable from the data repository The Open Science Framework (OSF) at https://doi.org/10.17605/OSF.IO/HKBSG or available as supplementary materials attached to the article.

## References

1. Hastrup LH, Simonsen E, Ibsen R, et al. Societal Costs of Schizophrenia in Denmark: A Nationwide Matched Controlled Study of Patients and Spouses Before and After Initial Diagnosis. Schizophrenia bulletin 2020;46(1):68–77. doi: 10.1093/schbul/sbz041 [published Online First: 2019/06/13]

2. Correll CU, Galling B, Pawar A, et al. Comparison of Early Intervention Services vs Treatment as Usual for Early-Phase Psychosis: A Systematic Review, Meta-analysis, and Meta-regression. JAMA Psychiatry 2018;75(6):555–65. doi: 10.1001/jamapsychiatry.2018.0623 [published Online First: 2018/05/26]

3. Friis S, Melle I, Johannessen JO, et al. Early Predictors of Ten-Year Course in First-Episode Psychosis. Psychiatric services (Washington, DC) 2016;67(4):438–43. doi: 10.1176/appi.ps.201400558

4. Kane JM, Agid O, Baldwin ML, et al. Clinical Guidance on the Identification and Management of Treatment-Resistant Schizophrenia. The Journal of clinical psychiatry 2019;80(2) doi: 10.4088/JCP.18com12123 [published Online First: 2019/03/07]

5. Elkis H. Treatment-resistant schizophrenia. The Psychiatric clinics of North America 2007;30(3):511-33. doi: 10.1016/j.psc.2007.04.001 [published Online First: 2007/08/28]

6. Howes OD, McCutcheon R, Agid O, et al. Treatment-Resistant Schizophrenia: Treatment Response and Resistance in Psychosis (TRRIP) Working Group Consensus Guidelines on Diagnosis and Terminology. The American journal of psychiatry 2017;174(3):216–29. doi: 10.1176/appi.ajp.2016.16050503

7. Huhn M, Nikolakopoulou A, Schneider-Thoma J, et al. Comparative efficacy and tolerability of 32 oral antipsychotics for the acute treatment of adults with multi-episode schizophrenia: a systematic review and network meta-analysis. The Lancet 2019;394(10202):939–51. doi: 10.1016/S0140-6736%2819%2931135-3

8. Kane JM, Agid O, Baldwin ML, et al. Clinical Guidance on the Identification and Management of Treatment-Resistant Schizophrenia. The journal of clinical psychiatry 2019;80(2) doi: 10.4088/JCP.18com12123

9. Nielsen J, Young C, Ifteni P, et al. Worldwide Differences in Regulations of Clozapine Use. CNS drugs 2016;30(2):149–61. doi: 10.1007/s40263-016-0311-1

10. Doyle R, Behan C, O’Keeffe D, et al. Clozapine Use in a Cohort of First-Episode Psychosis. Journal of clinical psychopharmacology 2017;37(5):512–17. doi: 10.1097/jcp.0000000000000734 [published Online First: 2017/06/27]

11. Howes OD, Vergunst F, Gee S, et al. Adherence to treatment guidelines in clinical practice: study of antipsychotic treatment prior to clozapine initiation. The British journal of psychiatry : the journal of mental science 2012;201(6):481–5. doi: 10.1192/bjp.bp.111.105833 [published Online First: 2012/09/08]

12. Bachmann CJ, Aagaard L, Bernardo M, et al. International trends in clozapine use: a study in 17 countries. Acta psychiatrica Scandinavica 2017;136(1):37–51. doi: 10.1111/acps.12742

13. Xiang Y-T, Wang C-Y, Si T-M, et al. Clozapine use in schizophrenia: findings of the Research on Asia Psychotropic Prescription (REAP) studies from 2001 to 2009. Australian and New Zealand journal of psychiatry 2011;45(11):968–75. doi: 10.3109/00048674.2011.607426

14. Stroup TS, Gerhard T, Crystal S, et al. Geographic and Clinical Variation in Clozapine Use in the United States. Psychiatric services (Washington, DC) 2014;65(2):186–92. doi: 10.1176/appi.ps.201300180

15. Gören JL, Meterko M, Williams S, et al. Antipsychotic Prescribing Pathways, Polypharmacy, and Clozapine Use in Treatment of Schizophrenia. Psychiatric Services 2013;64(6):527–33. doi: 10.1176/appi.ps.002022012

16. Bogers JP, Schulte PF, Van Dijk D, et al. Clozapine Underutilization in the Treatment of Schizophrenia: How Can Clozapine Prescription Rates Be Improved? J Clin Psychopharmacol 2016;36(2):109–11. doi: 10.1097/jcp.0000000000000478 [published Online First: 2016/02/13]

17. Thien K, O’Donoghue B. Delays and barriers to the commencement of clozapine in eligible people with a psychotic disorder: A literature review. Early Interv Psychiatry 2019;13(1):18–23. doi: 10.1111/eip.12683 [published Online First: 2018/07/10]

18. John AP, Ko EKF, Dominic A. Delayed Initiation of Clozapine Continues to Be a Substantial Clinical Concern. Can J Psychiatry 2018;63(8):526–31. doi: 10.1177/0706743718772522 [published Online First: 2018/04/24]

19. Baig AI, Bazargan-Hejazi S, Ebrahim G, et al. Clozapine prescribing barriers in the management of treatment-resistant schizophrenia: A systematic review. Medicine 2021;100(45):e27694–e94.

20. Farooq S, Choudry A, Cohen D, et al. Barriers to using clozapine in treatment-resistant schizophrenia: systematic review. BJPsych Bull 2019;43(1):8–16. doi: 10.1192/bjb.2018.67 [published Online First: 2018/09/29]

21. Verdoux H, Quiles C, Bachmann CJ, et al. Prescriber and institutional barriers and facilitators of clozapine use: A systematic review. Schizophrenia research 2018;201:10–19. doi: 10.1016/j.schres.2018.05.046

22. Taylor D, Shapland L, Laverick G, et al. Clozapine - A survey of patient perceptions. Psychiatric Bulletin 2000;24(12):450–52.

23. Bogers JP, Bui H, Herruer M, et al. Capillary compared to venous blood sampling in clozapine treatment: patients’ and healthcare practitioners’ experiences with a point-of-care device. European neuropsychopharmacology 2015;25(3):319–24. doi: 10.1016/j.euroneuro.2014.11.022

24. Nielsen J, Thode D, Stenager E, et al. Hematological clozapine monitoring with a point-of-care device: a randomized cross-over trial. European neuropsychopharmacology 2012;22(6):401–05. doi: 10.1016/j.euroneuro.2011.10.001

25. Hodge K, Jespersen S. Side-effects and treatment with clozapine: a comparison between the views of consumers and their clinicians. International Journal of Mental Health Nursing 2008;17(1):2–8. doi: 10.1111/j.1447-0349.2007.00506.x

26. Michelle Iris J, Ole Jakob S, Stephen Fitzgerald A, et al. Patients’ and psychiatrists’ perspectives on clozapine treatment—a scoping review protocol. BMJ Open 2021;11(10):e054308. doi: 10.1136/bmjopen-2021-054308

27. Parkes S, Mantell B, Oloyede E, et al. Patients’ Experiences of Clozapine for Treatment-Resistant Schizophrenia: A Systematic Review. Schizophrenia Bulletin Open 2022;3(1) doi: 10.1093/schizbullopen/sgac042

28. Grover S, Naskar C. Patient and caregivers perspective about clozapine: A systematic review. Schizophr Res 2023 doi: 10.1016/j.schres.2023.06.005 [published Online First: 2023/06/30]

29. Kim JH, Kim SY, Ahn YM, et al. Subjective response to clozapine and risperidone treatment in outpatients with schizophrenia. Progress in Neuro-Psychopharmacology and Biological Psychiatry 2006;30(2):301–05.

30. Angermeyer MC, Loffler W, Muller P, et al. Patients’ and relatives’ assessment of clozapine treatment. Psychological Medicine 2001;31(3):509–17. doi: 10.1017/s0033291701003749

31. Murphy K, Coombes I, McMillan S, et al. Clozapine and shared care: the consumer experience. Australian Journal of Primary Health 2018;24(6):455–62. doi: 10.1071/PY18055

32. Qurashi I, Stephenson P, Chu S, et al. An evaluation of subjective experiences, effects and overall satisfaction with clozapine treatment in a UK forensic service. Therapeutic advances in psychopharmacology 2015;5(3):146–50.

33. Sharma S, Kopelovich SL, Janjua AU, et al. Cluster analysis of clozapine consumer perspectives and comparison to consumers on other antipsychotics. Schizophrenia Bulletin Open 2021;2(1):sgab043.

34. Siskind DJ, Harris M, Phillipou A, et al. Clozapine users in Australia: Their characteristics and experiences of care based on data from the 2010 National Survey of High Impact Psychosis. Epidemiology and Psychiatric Sciences 2017;26(3):325–37. doi: 10.1017/S2045796016000305

35. Sloan D, Hutchinson L, O’Boyle J. Client satisfaction in a clozapine clinic. European Psychiatry: the Journal of the Association of European Psychiatrists 1997;12(7):373.

36. Takeuchi H, Borlido C, Sanches M, et al. Adherence to clozapine vs. other antipsychotics in schizophrenia. Acta psychiatrica Scandinavica 2020;142(2):87–95. doi: 10.1111/acps.13208

37. Verma M, Grover S, Chakrabarti S, et al. Attitude towards and experience with clozapine of patients and their caregivers after three months of starting of clozapine. Nordic Journal of Psychiatry 2020

38. Waserman J, Criollo M. Subjective experiences of clozapine treatment by patients with chronic schizophrenia. Psychiatric Services 2000;51(5):666–68. doi: 10.1176/appi.ps.51.5.666

39. Wolfson PM, Paton C. Clozapine audit: what do patients and relatives think? Journal of Mental Health 1996;5(3):267–73. doi: 10.1080/09638239650036938

40. Srour A, Eltorki Y, Malik H, et al. Patients’ and primary carers’ views on clozapine treatment for schizophrenia: A cross-sectional study in Qatar. Saudi Pharmaceutical Journal 2023;31(2):214–21.

41. Gee SH, Shergill SS, Taylor DM. Patient attitudes to clozapine initiation. International clinical psychopharmacology 2017;32(6):337–42.

42. Lewis SW, Barnes TR, Davies L, et al. Randomized controlled trial of effect of prescription of clozapine versus other second-generation antipsychotic drugs in resistant schizophrenia. Schizophrenia Bulletin 2006;32(4):715–23. doi: 10.1093/schbul/sbj067

43. Li Q, Xiang Y-T, Su Y-A, et al. Clozapine in schizophrenia and its association with treatment satisfaction and quality of life: Findings of the three national surveys on use of psychotropic medications in China (2002-2012). Schizophrenia Research 2015;168(1/2):523–29. doi: 10.1016/j.schres.2015.07.048

44. Sowerby C, Taylor D. Cross-sector user and provider perceptions on experiences of shared-care clozapine: A qualitative study. BMJ Open 2017;7(9):581. doi: 10.1136/bmjopen-2017-017183

45. Arksey H, O’Malley L. Scoping studies: towards a methodological framework. International journal of social research methodology 2005;8(1):19–32. doi: 10.1080/1364557032000119616

46. Tricco AC, Lillie E, Zarin W, et al. PRISMA Extension for Scoping Reviews (PRISMA-ScR): Checklist and Explanation. Annals of internal medicine 2018;169(7):467–73. doi: 10.7326/M18-0850

47. Peters MDJ, Godfrey CM, Khalil H, et al. Guidance for conducting systematic scoping reviews. International journal of evidence-based healthcare 2015;13(3):141–46. doi: 10.1097/XEB.0000000000000050

48. Peters MDJ, Marnie C, Tricco AC, et al. Updated methodological guidance for the conduct of scoping reviews. JBI evidence synthesis 2020;18(10):2119–26. doi: 10.11124/JBIES-20-00167

49. Levac D, Colquhoun H, O’Brien KK. Scoping studies: advancing the methodology. Implementation science : IS 2010;5(1):69–69. doi: 10.1186/1748-5908-5-69

50. Patients’ and psychiatrists’ perspectives on clozapine treatment - a scoping review The Open Science Framework (OSF): The Center for Open Science (COS); 2021 [Available from: https://osf.io/5k4s3/?view_only=36a0c7fb1e694c18a9b5cef47cd77227.

51. Bramer W, Rethlefsen ML, Kleijnen J, et al. Optimal database combinations for literature searches in systematic reviews: A prospective exploratory study. Systematic reviews 2017;6(1):245–45. doi: 10.1186/s13643-0170644-y

52. EndNote [program]. EndNote X9 version. Philadelphia, PA: Clarivate, 2013.

53. Covidence systematic review software [3 program]. Melbourne, Australia: Veritas Health Innovation.

54. Microsoft Excel [program]. 2019 (16.0) version, 2018.

55. Hsieh HF, Shannon SE. Three approaches to qualitative content analysis. Qual Health Res 2005;15(9):1277–88. doi: 10.1177/1049732305276687 [published Online First: 2005/10/06]

56. Agarwal A, Singla N, Grover S. Patient’s perspective of Clozapine. Indian Journal of Psychiatry 2017;59(6):S167–S67.

57. Waterreus A, Morgan VA, Castle D, et al. Medication for psychosis – consumption and consequences: The second Australian national survey of psychosis. Australian & New Zealand Journal of Psychiatry 2012;46(8):762–73. doi: 10.1177/0004867412450471

58. Voruganti L, Cortese L, Oyewumi L, et al. Comparative evaluation of conventional and novel antipsychotic drugs with reference to their subjective tolerability, side-effect profile and impact on quality of life. Schizophrenia Research 2000;43(2-3):135–45.

59. Tilley S, Chambers M. Does patient education enhance compliance with clozapine? A preliminary investigation. Journal of Psychiatric & Mental Health Nursing (Wiley-Blackwell) 2000;7(3):285.

60. Balestrieri M, Di Sciascio G, Isola M, et al. Drug attitude and subjective well-being in antipsychotic treatment monotherapy in real-world settings. Epidemiologia e Psichiatria Sociale 2009;18(2):114–18.

61. Castle D, Morgan V, Jablensky A. Antipsychotic use in Australia: the patients’ perspective. Australian & New Zealand Journal of Psychiatry 2002;36(5):633–41. doi: 10.1046/j.1440-1614.2002.01037.x

62. Paruk S, Roojee SA. Attitude and knowledge towards clozapine among outpatients prescribed clozapine in Durban, KwaZulu-Natal. South African Journal of Psychiatry 2018;24(1):1–1. doi: 10.4102/sajpsychiatry.v24i0.1316

63. Nordon C, Rouillon F, Barry C, et al. Determinants of treatment satisfaction of schizophrenia patients: results from the ESPASS study. Schizophrenia Research 2012;139(1-3):211–17. doi: 10.1016/j.schres.2012.05.024

64. Munro J, Laverick G, Shapland L, et al. A clozapine patient questionnaire: An insight into the patient’s perspective. Schizophrenia Research 2000;41(1):184–84.

65. McAllister M, Chatterton R. Clozapine: exploring clients’ experiences of treatment. The Australian and New Zealand journal of mental health nursing 1996;5(3):136–42.

66. Man WH, Colen-de Koning J, Schulte P, et al. Clozapine-induced hypersalivation: the association between quantification, perceived burden and treatment satisfaction reported by patients. Therapeutic advances in psychopharmacology 2017;7(8-9):209–10.

67. Dickens GL, Frogley C, Mason F, et al. Experiences of women in secure care who have been prescribed clozapine for borderline personality disorder. Borderline Personality Disorder and Emotion Dysregulation 2016;3:12.

68. Leijala J, Kampman O, Suvisaari J, et al. Daily functioning and symptom factors contributing to attitudes toward antipsychotic treatment and treatment adherence in outpatients with schizophrenia spectrum disorders. BMC psychiatry 2021;21(1)

69. Kuroda N, Sun S, Lin CK, et al. Attitudes toward taking medication among outpatients with schizophrenia: Cross-national comparison between Tokyo and Beijing. Environmental Health and Preventive Medicine 2008;13(5):288–95.

70. Krzystanek M, Krysta K, Janas-Kozik M, et al. Risk factors for noncompliance with antipsychotic medication in long-term treated chronic schizophrenia patients. Psychiatria Danubina 2019;31:S543–S48.

71. Freudenreich O, Cather C, Evins AE, et al. Attitudes of schizophrenia outpatients toward psychiatric medications: relationship to clinical variables and insight. The Journal of clinical psychiatry 2004;65(10):1372–76.

72. Kaar SJ, Gobjila C, Butler E, et al. Making decisions about antipsychotics: A qualitative study of patient experience and the development of a decision aid. BMC psychiatry 2019;19 (1)

73. Jenkins JH, Strauss ME, Carpenter EA, et al. Subjective experience of recovery from schizophrenia-related disorders and atypical antipsychotics. International Journal of Social Psychiatry 2005;51(3):211–27.

74. Frogley C, Anagnostakis K, Mitchell S, et al. A case series of clozapine for borderline personality disorder. Annals of clinical psychiatry 2013;25(2):125–34.

75. Gray R, Smedley N, Miller K, et al. Research in brief. Health education needs of people with schizophrenia taking clozapine. Journal of Clinical Nursing (Wiley-Blackwell) 1996;5(5):333–34. doi: 10.1111/jocn.1996.5.5.333

76. Garcia Cabeza I, Sanz Amador M, Arango Lopez C, et al. Subjective response to antipsychotics in schizophrenic patients: Clinical implications and related factors. Schizophrenia Research 2000;41(2):349–55.

77. Bowen J. Obtaining consent for treatment with clozapine. Psychiatric Bulletin 1992;16(4):239–40.

78. Naber D, Riedel M, Klimke A, et al. Randomized double blind comparison of olanzapine vs. clozapine on subjective wellbeing and clinical outcome in patients with schizophrenia. Acta psychiatrica Scandinavica 2005;111(2):106–15.

79. Lindstrom LH. The effect of long-term treatment with clozapine in schizophrenia: A retrospective study in 96 patients treated with clozapine for up to 13 years. Acta psychiatrica Scandinavica 1988;77(5):524–29.

80. Blagden S, Beenstock J, Auld N, et al. A qualitative exploration of the barriers to and facilitators of clozapine monitoring in a secure psychiatric setting. BJPsych Bulletin 2020:1–7.

81. De La Salle B. Point-of-care testing in clinical practice: Applications in the mental health setting. International Journal of Laboratory Hematology 2010;32:55–56.

82. Kelly DL, Ponomareva OY, Mackowick M, et al. Feasibility and patient-reported satisfaction using a novel point-of-care fingerstick method for monitoring absolute neutrophil count for clozapine. Annals of clinical psychiatry : official journal of the American Academy of Clinical Psychiatrists 2021;33(2):116–23.

83. Sowerby C, Taylor D. Cross-sector user and provider perceptions on experiences of shared-care clozapine: A qualitative study. BMJ Open 2017;7(9)

84. Takeuchi I, Hanya M, Uno J, et al. A questionnaire-based study of the views of schizophrenia patients and psychiatric healthcare professionals in Japan about the side effects of clozapine. Clinical Psychopharmacology and Neuroscience 2016;14(3):286–94.

85. Taylor D, Sutton J, Family H. Evaluating the Pharmacist Provision of Clozapine Services: University of Bath, 2011:23.

86. Apiquian R, Fresan A, de la Fuente-Sandoval C, et al. Survey on schizophrenia treatment in Mexico: Perception and antipsychotic prescription patterns. BMC psychiatry 2004;4 (no pagination)

87. Bahji A, Bajaj N. Attitudes to antipsychotics: a multi-site survey of Canadian psychiatry residents. Journal of Mental Health Training, Education & Practice 2018;13(6):318–38. doi: 10.1108/JMHTEP-03-2018-0019

88. Bleakley S, Olofinjana O, Taylor D. Which antipsychotics would mental health professionals take themselves? Psychiatric Bulletin 2007;31(3):94–96.

89. Chane-Kene A, Plassart F, Chauvel O, et al. A new breath for clozapine. European Journal of Hospital Pharmacy 2019;26 (Supplement 1):A150.

90. Chow EWC, Collins EJ, Nuttall SE, et al. Clinical use of clozapine in a major urban setting: One year experience. Journal of Psychiatry and Neuroscience 1995;20(2):133–40.

91. Correll CU, Brevig T, Brain C. Patient characteristics, burden and pharmacotherapy of treatment-resistant schizophrenia: results from a survey of 204 US psychiatrists. BMC psychiatry 2019;19(1)

92. Cotes RO, Janjua AU, Broussard B, et al. A Comparison of Attitudes, Comfort, and Knowledge of Clozapine Among Two Diverse Samples of US Psychiatrists. Community mental health journal 2021;29

93. Daod E, Krivoy A, Shoval G, et al. Psychiatrists’ attitude towards the use of clozapine in the treatment of refractory schizophrenia: A nationwide survey. Psychiatry research 2019;275:155–61.

94. De Hert M, De Beugher A, Sweers K, et al. Knowledge of Psychiatric Nurses About the Potentially Lethal Side-Effects of Clozapine. Archives of Psychiatric Nursing 2016;30 (1):79–83. doi: 10.1016/j.apnu.2015.09.003

95. Dvalishvili M, Miller BJ, Surya S. Comfort Level and Perceived Barriers to Clozapine Use: Survey of General Psychiatry Residents. Academic psychiatry : the journal of the American Association of Directors of Psychiatric Residency Training and the Association for Academic Psychiatry 2021;12

96. El Hayek S, Noufi P, Beayno A, et al. Prescribing Clozapine in the MENA Region: The Perspective and Practice of Psychiatrists. Arab Journal of Psychiatry 2021;32(1):64–78.

97. Feakins M, Odejayi G, Groll D, et al. Are we making the most of clozapine? European Psychiatry 2015;30:1685.

98. Gan Y-L, O’Reilly CL, O’Reilly CL. Community pharmacists’ attitudes and opinions towards supplying clozapine. International Journal of Clinical Pharmacy 2018;40(5):1116–30. doi: 10.1007/s11096-018-0676-y

99. Gee S, Vergunst F, Howes O, et al. Practitioner attitudes to clozapine initiation. Acta psychiatrica Scandinavica 2014;130(1):16–24. doi: 10.1111/acps.12193

100. Gören JL, Rose AJ, Engle RL, et al. Organizational Characteristics of Veterans Affairs Clinics With High and Low Utilization of Clozapine. Psychiatric Services 2016;67(11):1189–96. doi: 10.1176/appi.ps.201500506

101. Grau-Lopez L, Szerman N, Torrens M, et al. Professional perception of clozapine use in patients with dual psychosis. Actas espanolas de psiquiatria 2020;48(3):99–105.

102. Grover S, Balachander S, Chakarabarti S, et al. Prescription practices and attitude of psychiatrists towards clozapine: A survey of psychiatrists from India. Asian Journal of Psychiatry 2015;18:57–65.

103. Haw C, Stubbs J. Medication for borderline personality disorder: A survey at a secure hospital. International Journal of Psychiatry in Clinical Practice 2011;15(4):270–74. doi: 10.3109/13651501.2011.590211

104. Henry R, Massey R, Morgan K, et al. Evaluation of the effectiveness and acceptability of intramuscular clozapine injection: Illustrative case series. BJPsych Bulletin 2020;44(6):239–43. doi: 10.1192/bjb.2020.6

105. Ismail D, Tounsi K, Zolezzi M, et al. A qualitative exploration of clozapine prescribing and monitoring practices in the Arabian Gulf countries. Asian Journal of Psychiatry 2019;39:93–97. doi: 10.1016/j.ajp.2018.12.011

106. Jauhar S, Guloksuz S, Gama Marques J, et al. Treatment choice in psychiatry? How would European trainees treat psychosis for their patients and themselves, and what influences decision-making. European Psychiatry Conference: 18th European Congress of Psychiatry Munich Germany Conference Publication: 2010;25(SUPPL. 1)

107. Ketter TA, Haupt DW, Ketter TA, et al. Perceptions of weight gain and bipolar pharmacotherapy: results of a 2005 survey of physicians in clinical practice. Current Medical Research & Opinion 2006;22(12):2345–53.

108. Knowles S-A, McMillan SS, Wheeler AJ. Consumer access to clozapine in Australia: how does this compare to New Zealand and the United Kingdom? Pharmacy Practice (1886-3655) 2016;14(2):1-9. doi: 10.18549/PharmPract.2016.02.722

109. Latas M, Stojkovic T, Ralic T, et al. Psychiatrists’ psychotropic drug prescription preferences for themselves or their family members. Psychiatria Danubina 2012;24(2):182–87.

110. Leung JG, Cusimano J, Gannon JM, et al. Addressing clozapine under-prescribing and barriers to initiation: A psychiatrist, advanced practice provider, and trainee survey. International clinical psychopharmacology 2019;34(5):247–56.

111. Lonnen J, McNeil L, Capek E, et al. A survey of the management of psychosis in Parkinson’s disease. Movement Disorders 2012;27:S23.

112. Love RC, Mackowick M, Carpenter D, et al. Expert consensus-based medication-use evaluation criteria for atypical antipsychotic drugs. American Journal of Health-System Pharmacy 2003;60(23):2455–70. doi: 10.1093/ajhp/60.23.2455

113. Maryan S, Harms M, McAllister E, et al. Comparison of clozapine monitoring and adverse event management in a psychiatrist-only and a clinical pharmacist-psychiatrist collaborative clinic. The Mental Health Clinician 2019;9(2):70–75.

114. Melse U, Kurz M, Fleischhacker WW. Antipsychotic maintenance treatment of schizophrenia patients: Is there a consensus? Schizophrenia Bulletin 1994;20(1):215–25.

115. Mishara AL, Orr B, Buckley P. Update on psychopharmacology. Staff perceptions of clozapine and their role in treatment: initial observations. Journal of Psychosocial Nursing & Mental Health Services 1995;33(10):44–47.

116. Moeller FG, Chen YW, Steinberg JL, et al. Risk factors for clozapine discontinuation among 805 patients in the VA Hospital System. Annals of clinical psychiatry 1995;7(4):167–73.

117. Moody BL, Eatmon CV. Perceived Barriers and Facilitators of Clozapine Use: A National Survey of Veterans Affairs Prescribers. Federal Practitioner 2019;36(Suppl 6):S22–S27.

118. Mortimer AM, Singh P, Shepherd CJ, et al. Clozapine for treatment-resistant schizophrenia: National Institute of Clinical Excellence (NICE) guidance in the real world. Clinical Schizophrenia and Related Psychoses 2010;4(1):49–55.

119. Nielsen J, Dahm M, Lublin H, et al. Psychiatrists’ attitude towards and knowledge of clozapine treatment. Journal of Psychopharmacology 2010;24(7):965–71. doi: 10.1177/0269881108100320

120. O’Brien A, Firn M. Clozapine initiation in the community. Psychiatric Bulletin 2002;26(9):339–41.

121. Okhuijsen-Pfeifer C, Cohen D, Bogers JPAM, et al. Differences between physicians’ and nurse practitioners’ viewpoints on reasons for clozapine underprescription. Brain and Behavior 2019;9(7)

122. Paranthaman R, Baldwin RC. Survey of clozapine use by consultant old age psychiatrists. Psychiatric Bulletin 2006;30(11):410–12.

123. Fricchione Parise V, Balletta G, Manna G. Clozapine utilisation in routine clinical practice: Golden opportunity or neglected orphan? Real-world outcomes. European neuropsychopharmacology 2013;23:S479–S80.

124. Paton C, Garcia JA, Brooke D. Use of atypical antipsychotics by consultant psychiatrists working in forensic settings. Psychiatric Bulletin 2002;26(5):172–74.

125. Péré JJ, Chaumet-Riffaud PD, Bourdeix I, et al. Clozapine (Leponex) in France. Encephale 1992;18 Spec No 3:427–32.

126. Pereira S, Beer D, Paton C. Enforcing treatment with clozapine: Survey of views and practice. Psychiatric Bulletin 1999;23(6):342–45.

127. Ur-Rahman R, Ansari MA, Khan AG, et al. Preferred antipsychotic by mental health professionals of Sindh and Balauchistan. Journal of the Liaquat University of Medical and Health Sciences 2010;9(2):95–100.

128. Ignjatovic Ristic D, Cohen D, Ristic I. Prescription attitudes and practices regarding clozapine among Serbian psychiatrists: results of a nationwide survey. Therapeutic advances in psychopharmacology 2021;11(no pagination)

129. Sayer M, Love R, Freudenreich O, et al. Improving clozapine use in the united states: A survey of barriers and solutions informing a workgroup to devise a national strategy. Schizophrenia Bulletin 2017;43 (Supplement 1):S252.

130. Shrivastava A, Shah N. Prescribing practices of clozapine in India: Results of a opinion survey of psychiatrists. Indian Journal of Psychiatry 2009;51(3):230–31.

131. Silveira ASD, Rocha D, Attux CRD, et al. Patterns of clozapine and other antipsychotics prescriptions in patients with treatment-resistant schizophrenia in community mental health centers in Sao Paulo, Brazil. Archives of Clinical Psychiatry 2015;42(6):165–70. doi: 10.1590/0101-60830000000069

132. Singh B, Hughes AJ, Roerig JL. Comfort Level and Barriers to the Appropriate Use of Clozapine: a Preliminary Survey of US Psychiatric Residents. Academic Psychiatry 2020;44(1):53–58. doi: 10.1007/s40596-019-01134-7

133. Sutton J, Family H, Scott J, et al. The influence of organisational climate on care of patients with schizophrenia: a qualitative analysis of health care professionals’ views. International Journal of Clinical Pharmacy 2016;38(2):344–52. doi: 10.1007/s11096-016-0247-z

134. Swinton M, Ahmed AG. Reasons for non-prescription of clozapine in treatment-resistant schizophrenia. Criminal Behaviour and Mental Health 1999;9(3):207–14.

135. Taylor M, Brown T. “Do unto others as…” - Which treatments do psychiatrists prefer? Results from a national survey. Scottish Medical Journal 2007;52(1):17–19.

136. Tungaraza TE, Farooq S. Clozapine prescribing in the UK: views and experience of consultant psychiatrists. Therapeutic advances in psychopharmacology 2015;5(2):88–96.

137. Udomratn P, Srisurapanont M. Impact on Thai psychiatrists of passive dissemination of a clinical practice guideline on prescribing attitudes in treatment-resistant schizophrenia. Neuropsychobiology 2002;45(4):186–90.

138. Wilson B, McMillan SS, Wheeler AJ. Implementing a clozapine supply service in Australian community pharmacies: Barriers and facilitators. Journal of Pharmaceutical Policy and Practice 2019;12(1)

139. van der Zalm YC, Schulte PF, Bogers J, et al. Delegating clozapine monitoring to advanced nurse practitioners: An exploratory, randomized study to assess the effect on prescription and its safety. Administration and Policy in Mental Health and Mental Health Services Research 2020;47(4):632–40.

140. Agid O, Remington G, Kapur S, et al. Early use of clozapine for poorly responding first-episode psychosis. Journal of clinical psychopharmacology 2007;27(4):369–73.

141. Atkinson JM, Douglas-Hall P, Fischetti C, et al. Outcome following clozapine discontinuation: A retrospective analysis. Journal of Clinical Psychiatry 2007;68(7):1027–30. doi: 10.4088/jcp.v68n0708

142. Baker M, White T. Life after clozapine. Medicine, Science and the Law 2004;44(3):217–21.

143. Black LL, Greenidge LL, Ehmann T, et al. A centralized system for monitoring clozapine use in British Columbia. Psychiatric Services 1996;47(1):81–83.

144. Casetta C, Oloyede E, Whiskey E, et al. A retrospective study of intramuscular clozapine prescription for treatment initiation and maintenance in treatment-resistant psychosis. British Journal of Psychiatry 2020;217(3):506–13. doi: 10.1192/bjp.2020.115

145. Cassano GB, Ciapparelli A, Villa M. Clozapine as a treatment tool: Only in resistant schizophrenic patients? European Psychiatry 1997;12(SUPPL. 5):347s–51s.

146. Chen EYH, Stubbs JH, Staley CJ. Pattern of clozapine use in 64 consecutive patients in a tertiary care hospital. Pharmaceutical Journal 1996;256(6892):683–85.

147. Chengappa KNR, Baker RW, Kreinbrook SB, et al. Clozapine use in female geriatric patients with psychoses. Journal of Geriatric Psychiatry and Neurology 1995;8(1):12–15.

148. Dalal B, Larkin E, Leese M, et al. Clozapine treatment of long-standing schizophrenia and serious violence: A two-year follow-up study of the first 50 patients treated with clozapine in Rampton high security hospital. Criminal Behaviour and Mental Health 1999;9(2):168–78.

149. Davis MC, Fuller MA, Strauss ME, et al. Discontinuation of clozapine: a 15-year naturalistic retrospective study of 320 patients. Acta psychiatrica Scandinavica 2014;130(1):30–39. doi: 10.1111/acps.12233 [published Online First: Davis MC, Fuller MA, Strauss ME, Konicki PE, Jaskiw GE. Discontinuation of clozapine: A 15-year naturalistic retrospective study of 320 patients.]

150. Drew LR, Griffiths KM, Hodgson DM. A five year follow-up study of the use of clozapine in community practice. Australian & New Zealand Journal of Psychiatry 2002;36(6):780–86. doi: 10.1046/j.1440-1614.2002.01091.x

151. Friedman JH, Goldstein S, Jacques C. Substituting clozapine for olanzapine in psychiatrically stable parkinson’s disease patients: Results of an open label pilot study. Clinical neuropharmacology 1998;21(5):285–88.

152. Gale E, Richardson CM, Vyas G, et al. Fifteen year follow up of clozapine treated patients with schizophrenia. Schizophrenia Bulletin 2013;39:S331.

153. Gonzalez C, Kodimela K, Poynton A. Clozapine initiation in crisis teams. British Journal of Medical Practitioners 2013;6(3)

154. Gee SH, Shergill SS, Taylor DM. Long-term follow-up of clozapine prescribing. Journal of Psychopharmacology 2018;32(5):552–58. doi: 10.1177/0269881118760666

155. Grace JJ, Szarowicz RS. The clozapine access project. Psychiatric services (Washington, DC) 2001;52(1):108.

156. Hack N, Fayad SM, Monari EH, et al. An eight-year clinic experience with clozapine use in a Parkinson’s disease clinic setting. Plos one 2014;9(3)

157. Klein C, Gordon J, Pollak L, et al. Clozapine in Parkinson’s disease psychosis: 5-year follow-up review. Clinical neuropharmacology 2003;26(1):8–11.

158. Krivoy A, Malka L, Fischel T, et al. Predictors of clozapine discontinuation in patients with schizophrenia. International clinical psychopharmacology 2011;26(6):311–15.

159. Law A, Croucher M. Prescribing trends and safety of clozapine in an older persons mental health population. International Psychogeriatrics 2019;31(12):1823–29.

160. Leclerc LD, Demers MF, Bardell A, et al. A Chart Audit Study of Clozapine Utilization in Early Psychosis. Journal of clinical psychopharmacology 2021;41(3):275–80.

161. Legge SE, Hamshere M, Hayes RD, et al. Reasons for discontinuing clozapine: A cohort study of patients commencing treatment. Schizophrenia Research 2016;174(1-3):113–19. doi: 10.1016/j.schres.2016.05.002

162. MacGillivray S, Cooper SJ, English B, et al. Predictors of discontinuation on clozapine: A population study. Irish Journal of Psychological Medicine 2003;20(4):115–18.

163. Macpherson R, Prasad Sarkar S, Medina-Galera JL, et al. Gloucester clozapine clinic. Psychiatric Bulletin 1998;22(5):300–02.

164. Martin A, O’Driscoll C, Samuels A. Clozapine use in a forensic population in a New South Wales prison hospital. Australian & New Zealand Journal of Psychiatry 2008;42(2):141–46. doi: 10.1080/00048670701787529

165. Mustafa FA, Burke JG, Abukmeil SS, et al. Schizophrenia past Clozapine: Reasons for Clozapine Discontinuation, Mortality, and Alternative Antipsychotic Prescribing. Pharmacopsychiatry 2014;45(1):11–14.

166. O’Connor DW, Sierakowski C, Chin LF, et al. The safety and tolerability of clozapine in aged patients: A retrospective clinical file review. World Journal of Biological Psychiatry 2010;11(6):788–91.

167. Ojo T, Abayomi O. Pattern of clozapine use among patients in a nigerian neuropsychiatric hospital. International journal of neuropsychopharmacology 2012;15:59.

168. Pai NB, Vella SC. Reason for clozapine cessation. Acta psychiatrica Scandinavica 2012;125(1):39–44.

169. Pfeiffer RF, Kang J, Graber B, et al. Clozapine for psychosis in Parkinson’s disease. Movement Disorders 1990;5(3):239–42.

170. Phaldessai S, Butler R, Radhakrishnan R, et al. Clozapine prescribing in older adults. International Journal of Geriatric Psychiatry 2019;34(7):1105–06.

171. Rascati KL, Rascati EJ. Use of clozapine in Texas state mental health facilities. American Journal of Hospital Pharmacy 1993;50(8):1663–66. doi: 10.1093/ajhp/50.8.1663

172. Hun Senol S, Gurcan G, Ertugrul A, et al. A major challenge for clinicians: Discussing rechallenge with clozapine through a case series. European neuropsychopharmacology 2017;27 (Supplement 4):S961.

173. Shaker A, Jones R. Clozapine discontinuation in early schizophrenia: a retrospective case note review of patients under an early intervention service. Therapeutic Advances in Psychopharmacology 2018;8(1):3–11. doi: 10.1177/2045125317741449

174. Tahnee B, Hesitha A, Arulmathy A, et al. The use of clozapine in a rural and remote region of Australia. Australasian Psychiatry 2021;29(2):134–38.

175. Taylor DM, Douglas-Hall P, Olofmjana B, et al. Reasons for discontinuing clozapine: Matched, case-control comparison with risperidone long-acting injection. British Journal of Psychiatry 2009;194(2):165–67.

176. Thalayasingam S, Alexander RT, Singh I. The use of clozapine in adults with intellectual disability. Journal of Intellectual Disability Research 2004;48(6):572–79. doi: 10.1111/j.1365-2788.2004.00626.x

177. Thien K, Bowtell M, Eaton S, et al. Clozapine use in early psychosis. Schizophrenia Research 2018;199:374–79. doi: 10.1016/j.schres.2018.02.054

178. Thomas AA, Friedman JH. Current use of clozapine in Parkinson disease and related disorders. Clinical neuropharmacology 2010;33(1):14–16.

179. Trosch RM, Friedman JH, Lannon MC, et al. Clozapine use in Parkinson’s disease: A retrospective analysis of a large multicentered clinical experience. Movement Disorders 1998;13(3):377–82.

180. Ucok A, Yagcioglu EA, Yildiz M, et al. Reasons for clozapine discontinuation in patients with treatment-resistant schizophrenia. Psychiatry Research 2019;275:149–54. doi: 10.1016/j.psychres.2019.01.110

181. Van Mechelen C, Uzair F. Audit of attendance of clozapine clinic two years after first audit. European Psychiatry Conference: 21st European Congress of Psychiatry, EPA 2013;28(SUPPL. 1)

182. Whiskey E, Wykes T, Duncan-McConnell D, et al. Continuation of clozapine treatment: Practice makes perfect. Psychiatric Bulletin 2003;27(6):211–13. doi: 10.1192/pb.27.6.211

183. Zito JM, Volavka J, Craig TJ, et al. Pharmacoepidemiology of clozapine in 202 inpatients with schizophrenia. Annals of Pharmacotherapy 1993;27(10):1262–69.

184. Bogers J, Cohen D. Venous compared to capillary blood sampling with a point-of care device in clozapine treatment: Patients’ preferences. European archives of psychiatry and clinical neuroscience 2015;265(1):S82.

185. Drew LR, Hodgson DM, Griffiths KM. Clozapine in community practice: a 3-year follow-up study in the Australian Capital Territory. Australian & New Zealand Journal of Psychiatry 1999;33(5):667–75. doi: 10.1080/j.1440-1614.1999.00631.x

186. Kalaria SN, Kelly DL. Development of point-of-care testing devices to improve clozapine prescribing habits and patient outcomes. Neuropsychiatric Disease and Treatment 2019;15:2365–70. doi: 10.2147/NDT.S216803

187. Kelly D, Glassman M, Mackowick M, et al. Satisfaction with using a novel fingerstick for absolute neutrophil count (ANC) at the point of treatment in patients treated with clozapine. Schizophrenia Bulletin 2020;46 (Supplement 1):S20–S21.

188. Kelly DL, Ben-Yoav H, Payne GF, et al. Blood draw barriers for treatment with clozapine and development of a point-of-care monitoring device. Clinical Schizophrenia and Related Psychoses 2018;12(1):23–30.

189. Kelly DL, Ben-Yoav H, Stock V, et al. Development of a lab-on-a-chip biosensor for clozapine monitoring. Neuropsychopharmacology 2013;38:S237–S38.

190. Lindstrom LH. A retrospective study on the long-term efficacy of clozapine in 96 schizophrenic and schizoaffective patients during a 13-year period. Psychopharmacology 1989;99(SUPPL.):S84–S86.

191. O’Donoghue B, Thien K, Mc Gorry P. Clozapine use within an early intervention for psychosis service. Australian and New Zealand journal of psychiatry 2019;53 (Supplement 1):91–92.

192. Roy MA, Demers MF, Crocker C, et al. The impact of clozapine use in early-intervention: A retrospective chart audit. Early intervention in psychiatry 2018;12 (Supplement 1):85.

193. Sowerby C, Taylor D. Exploring the perceptions and experiences of people who use and those that provide a shared care clozapine service. International Journal of Pharmacy Practice 2015;23:71–72.

194. Sutton J, Taylor DA, Dawson HE. Pharmacist prescribing in clozapine clinics. International Journal of Pharmacy Practice 2010;18:25.

195. Taylor D, Sutton J, Dawson H. User and staff perspectives of clozapine clinic services. International Journal of Pharmacy Practice 2010;18:72.

196. Jakobsen MI, Julie P Shaug, Ole J Storebø, et al. (S2) Individual study data (included studies); Patients’ and Clinicians’ Perspectives on Clozapine Treatment - a Scoping Review. 2024 doi: 10.17605/OSF.IO/HKBSG

197. Legge SE, Hamshere M, Hayes RD, et al. Reasons for discontinuing clozapine: A cohort study of patients commencing treatment. Schizophrenia research 2016;174(1-3):113–19. doi: 10.1016/j.schres.2016.05.002

198. Pai NB, Vella S. Reason for clozapine cessation. Acta Psychiatrica Scandinavica 2012;125(1):39–44.

199. Griffiths K, MacCabe J, Egerton A. A systematic review and metaanalysis of clinical variables associated with response to clozapine in treatment resistant schizophrenia. Schizophrenia Bulletin 2020;46(Supplement 1):S217.

200. Jakobsen MI, Austin SF, Storebø OJ, et al. Non-prescribing of clozapine for outpatients with schizophrenia in real-world settings: The clinicians’ perspectives. Schizophrenia 2023;9(1):91.

201. Southern J, Elliott P, Maidment I. What are patients’ experiences of discontinuing clozapine and how does this impact their views on subsequent treatment? BMC psychiatry 2023;23(1):1–14.

202. Oloyede E, Dunnett D, Taylor D, et al. The lived experience of clozapine discontinuation in patients and carers following suspected clozapine-induced neutropenia. BMC psychiatry 2023;23(1):1–7.

203. Rezaie L, Nazari A, Safari-Faramani R, et al. Iranian psychiatrists’ attitude towards clozapine use for patients with treatment-resistant schizophrenia: a nationwide survey. BMC psychiatry 2022;22(1):1–10.

204. Zheng S, Lee J, Chan SKW. Utility and barriers to clozapine use: a joint study of clinicians’ attitudes from Singapore and Hong Kong. The Journal of Clinical Psychiatry 2022;83(4):41090.

205. Rezaie L, Nazari A, Khazaie H. Exploration of the Barriers to Clozapine Prescribing in Patients with Treatment-Resistant Schizophrenia: A Qualitative Study. Journal of psychosocial rehabilitation and mental health 2023;10(1):45–53.

206. Oloyede E, Blackman G, Mantell B, et al. What are the barriers and facilitators of clozapine use in early psychosis? A survey of UK early intervention clinicians. Schizophrenia 2023;9(1):26.

207. Gangadharan D, Tirupati S. A qualitative study of clinicians’ perspectives on reasons for delays in clozapine initiation. Australasian Psychiatry 2023:10398562231177824.

208. Brugue O, Gonzalez M, Moreno L, et al. Clinician’s attitude towards clozapine prescription. European Psychiatry 2023;66(S1):S1013–S13.

209. Oloyede E, Mantell B, Williams J, et al. Clozapine for treatment resistance in early psychosis: a survey of UK clinicians’ training, knowledge and confidence. Therapeutic Advances in Psychopharmacology 2022;12:20451253221141222.

210. Torrens M, Basurte I, Vega P, et al. Professional perception of clozapine use in patients with dual psychosis. Actas Esp Psiquiatr 2020;48(3):99–105.

211. Verma M, Grover S, Chakrabarti S. Effectiveness of clozapine on quality of life and functioning in patients with treatment-resistant schizophrenia. Nordic Journal of Psychiatry 2021;75(2):135–44.

212. Grover S, Naskar C, Chakrabarti S. Experience with and attitude toward clozapine use among patients receiving clozapine on long term and their caregivers. Indian Journal of Psychiatry 2023;65(11):1165–75.

213. Butler E, Pillinger T, Brown K, et al. Real-world clinical and cost-effectiveness of community clozapine initiation: mirror cohort study. The British Journal of Psychiatry 2022;221(6):740–47.

214. Rowntree R, Murray S, Fanning F, et al. Clozapine use–has practice changed? Journal of Psychopharmacology 2020;34(5):567–73.

215. Jakobsen MI, Schaug JP, Nielsen J, et al. Antipsychotic prescribing practices for outpatients with schizophrenia and reasons for non-clozapine treatment - Data from a Danish quality assessment audit. Nordic Journal of Psychiatry 2023:1–10. doi: 10.1080/08039488.2022.2160878

